# The phenotypic spectrum of terminal and subterminal 6p deletions based on a social media-derived cohort and literature review

**DOI:** 10.1101/2022.11.08.22282033

**Authors:** Eleana Rraku, Wilhelmina S. Kerstjens-Frederikse, Morris A. Swertz, Trijnie Dijkhuizen, Conny M. A. van Ravenswaaij-Arts, Aafke Engwerda

**Author notes:** Author to whom correspondence should be addressed: Conny M.A. van Ravenswaaij-Arts, Department of Genetics, University Medical Centre Groningen, PO Box 30.001, 9700 RB Groningen, The Netherlands.

## Abstract

**Background:** Terminal 6p deletions are rare, and information on their clinical consequences is scarce, which impedes optimal management and follow-up by clinicians. The parent-driven Chromosome 6 Project collaborates with families of affected children worldwide to better understand the clinical effects of chromosome 6 aberrations and to support clinical guidance. A microarray report is required for participation, and detailed phenotype information is collected directly from parents through a multilingual web-based questionnaire. Information collected from parents is then combined with case data from literature reports. Here, we present our findings on 13 newly identified patients and 46 literature cases with genotypically well-characterised terminal and subterminal 6p deletions. We provide phenotype descriptions for both the whole group and for subgroups based on deletion size.

**Results:** The total group shared a common phenotype characterised by ocular anterior segment dysgenesis, vision problems, brain malformations, congenital defects of the cardiac septa and valves, mild to moderate hearing impairment, eye movement abnormalities, hypotonia, mild developmental delay and dysmorphic features. These characteristics were observed in all subgroups, confirming a dominant role for *FOXC1*, one of the most distally deleted genes. Additional characteristics were seen in individuals with terminal deletions exceeding 4.02 Mb, namely complex heart defects, corpus callosum abnormalities, kidney abnormalities and orofacial clefting. Some of these additional features may be related to the loss of other genes in the terminal 6p region, such as *RREB1* for the cardiac phenotypes and *TUBB2A* and *TUBB2B* for the cerebral phenotypes. In the newly identified patients, we observed previously unreported features including gastrointestinal problems, neurological abnormalities, balance problems and sleep disturbances.

**Conclusions:** We present an overview of the phenotypic characteristics observed in terminal and subterminal 6p deletions. This reveals a common phenotype that can be highly attributable to haploinsufficiency of *FOXC1*, with a possible additional effect of other genes in the 6p25 region. We also delineate the developmental abilities of affected individuals and report on previously unrecognised features, showing the added benefit of collecting information directly from parents. Based on our overview, we provide recommendations for clinical surveillance to support clinicians, patients and families.

## Background

Deletions including the end of the short arm of chromosome 6, terminal 6p, are rare and often result in variable phenotypes. In 2005, Lin et al. described the clinical features of 4 newly identified individuals and 18 literature reports of isolated terminal 6p deletions [1]. The authors stated that terminal 6p deletions result in a recognisable phenotype with commonly observed clinical features: ocular anterior chamber abnormalities, Dandy-Walker malformation, hearing impairment, cardiac anomalies, developmental delay, and a number of dysmorphic features, namely hypertelorism, down-slanting palpebral fissures, a flat nasal bridge and mid-face hypoplasia. This set of clinical features constitutes chromosome 6pter-p24 deletion syndrome (MIM#612582), which can be caused by terminal or subterminal interstitial 6pter-p24 deletions and shows high phenotypic variability [2–4]. The gene *FOXC1* (forkhead box c1, MIM*601090), located on 6p25.3, is considered the most influential gene for the 6p terminal deletion phenotype [2,5,6]. The largest review of individuals with 6pter-p24 deletion syndrome was published by de Vos et al. in 2017 [2], who described 69 patients with both terminal and subterminal deletions, including one new patient. However, de Vos et al. made no distinction between terminal and interstitial 6p25 deletions in their description of the phenotype. Most cases reviewed by Lin et al. and de Vos et al. were diagnosed using conventional cytogenetic techniques, and these authors attributed the phenotypic variability to differences in the deletion breakpoints, a theory also supported by others [7,8]. This phenotypic variability impedes optimal management and guidance of affected individuals by clinicians and leads to uncertainty for parents about their child’s future.

To help healthcare professionals and parents gain a better understanding of the clinical effects of chromosome 6 aberrations, the Chromosome 6 Project collaborates with parents of affected individuals worldwide via social media. Research on rare chromosome disorders is often limited by the small number of patients worldwide, and healthcare professionals often rely on information from literature reports and online databases that provide restricted and often incomplete information. With the widespread use of information technology and half the world’s population now using online social media platforms [9], these platforms have become a powerful means for researchers to engage with and recruit large numbers of participants and a forum to provide families and professionals with more detailed information.

In this study, we collected detailed phenotype and genotype information from 13 newly identified individuals with a terminal or subterminal 6p deletion and supplemented it with data from 46 literature cases. A microarray report was available for all 59 individuals, making this the largest cohort of genotypically well-characterised (sub)terminal 6p deletions to date. We used this data to develop a detailed phenotype description of (sub)terminal 6p deletions that also accounts for differences in deletion size. This work confirms the dominant role of *FOXC1* in the 6p terminal deletion phenotype, but we also discuss possible roles for other genes in some of the observed phenotypic differences. Most importantly, we provide recommendations for clinical surveillance based on the collected information.

## Methods

Phenotype and genotype data was collected for constitutional terminal and subterminal 6p deletion patients from the Chromosome 6 Project (called the “parent cohort” from here on) and from published literature reports (our “literature cohort”). Data collection and analysis were performed as described previously [10]. Terminal deletions were defined as deletions comprising the most distally located gene on chromosome 6p, *DUSP22* (dual-specificity phosphatase 22, MIM*616778). Subterminal deletions were defined as deletions that did not include *DUSP22* but did include at least part of the *FOXC1* gene given its prominent role in the 6p terminal deletion phenotype. We excluded individuals with intragenic *FOXC1* deletions and those with additional chromosomal aberrations with an expected phenotypic effect. We also excluded individuals with terminal 6p deletions from the literature cohort if their aberrations were benign according to the Database of Genomic Variants (DGV) (http://dgv.tcag.ca/dgv/app/home) [11].

### The parent cohort of patients recruited via the Chromosome 6 Project

Patients or their legal representatives were notified about the study via the Chromosome 6 Facebook support group, Twitter (@C6study) and the project’s website (www.chromosome6.org) [12]. To participate in the project, an official microarray report had to be uploaded during the sign-up procedure. The microarray analyses were performed in diagnostic laboratories using different platforms, and the results were converted to GRCh37/hg19 using the UCSC LiftOver Tool (https://genome.ucsc.edu) [13]. Phenotype data was collected directly from patients or their legal representatives, who were mostly parents, via the online multilingual Chromosome 6 Questionnaire, which parents could access through a personal account. The questionnaire, which was constructed using the MOLGENIS toolkit [14,15], consists of 132 closed-end questions about pregnancy, congenital abnormalities, dysmorphic features, development, behaviour and health-related problems. Participants were also approached to fill out a supplemental questionnaire about characteristics often reported in literature cases that were not yet part of the Chromosome 6 Questionnaire.

The phenotype and genotype data collected is stored in a secured database. Consent for publication was obtained for all participants. Consent for the use of photographs was optional. Data collected from individuals in the parent cohort was submitted to the DECIPHER database [16] (https://www.deciphergenomics.org).

### Literature cohort

We searched PubMed for previously reported patients with a terminal or subterminal 6p deletion using the search terms: chromosome 6p, 6p deletion, monosomy 6p and 6p*. Cases were further selected based on the abovementioned criteria and the availability of a microarray result and a detailed phenotype description. The reference lists of included case reports were checked for additional relevant cases. Phenotype data was extracted from the reports using the Chromosome 6 Questionnaire to ensure the homogeneity of the data collected.

### Data analysis

All clinical and behavioural characteristics were classified as present, absent, or unknown and are presented as present/known. Percentages are only given if the presence of a feature is known in at least 2/3rds of the individuals, and they are presented as a range with the minimum calculated as present/total number of cases × 100 and the maximum as (present + unknown)/total number of cases × 100. Development (intelligence quotient (IQ)) was categorised as normal (>85), borderline (70–85), mild (50–70), moderate (30–50), or severe (<30) delay. This was based on formal IQ test scores or, if these were not available, the mean of the developmental quotients for the milestones ‘walking independently’ and ‘using two-word sentences’. The developmental quotients were calculated as the 50^th^ centile of the population age of achievement for that milestone/the age of achievement in the participant × 100. The 50^th^ centiles of population age of achievement for the milestones ‘walking independently’ and ‘using two-word sentences’ are 12 [17] and 21 [18] months, respectively.

For all genes within the terminal 6p region (6p25.3p24.3), haploinsufficiency (HI) and loss-of-function intolerance (pLI) scores were extracted from DECIPHER (https://www.deciphergenomics.org) [16] in January 2021 (Table S2, Additional File 2). These scores are used to predict the HI-effect of a gene. They are derived from models that consider differences between known haploinsufficient and haplosufficient genes (HI score) [19] and protein-truncating variants seen in large-scale reference datasets (pLI score) [20]. Genes with an HI score between 0– 10% or a pLI score ≥0.9 are predicted to exhibit HI and thus to have a clinical effect when haploinsufficient. Therefore, they are called HI-genes. When applicable, we discuss the role of specific genes in the phenotype in the context of the observed differences in clinical characteristics.

Phenotype features are described for the whole group and for six subgroups. We also report the differences in the presence of clinical features between the parent and literature cohorts. Patients with a terminal deletion were divided into five subgroups (A–E) based on the presence or absence of certain HI-genes within the deletion (Figure 1): subgroup A – terminal deletion not including *FOXC1* (breakpoint distal to 1.61 Mb), subgroup B – terminal deletions including *FOXC1* but not *TUBB2A* (breakpoint between 1.61 and 3.15 Mb), subgroup C – terminal deletions including *TUBB2A* but not *PRPF4B* (breakpoint between 3.15 and 4.02 Mb), subgroup D – terminal deletions including *PRPF4B* but not *RREB1* (breakpoint between 4.02 and 7.11 Mb) and subgroup E – terminal deletions including *RREB1* (breakpoint proximal to 7.11 Mb). Subterminal deletions (distal breakpoint proximal to 0.35 Mb and including at least part of *FOXC1*) were assigned to subgroup S.

**Fig. 1.**
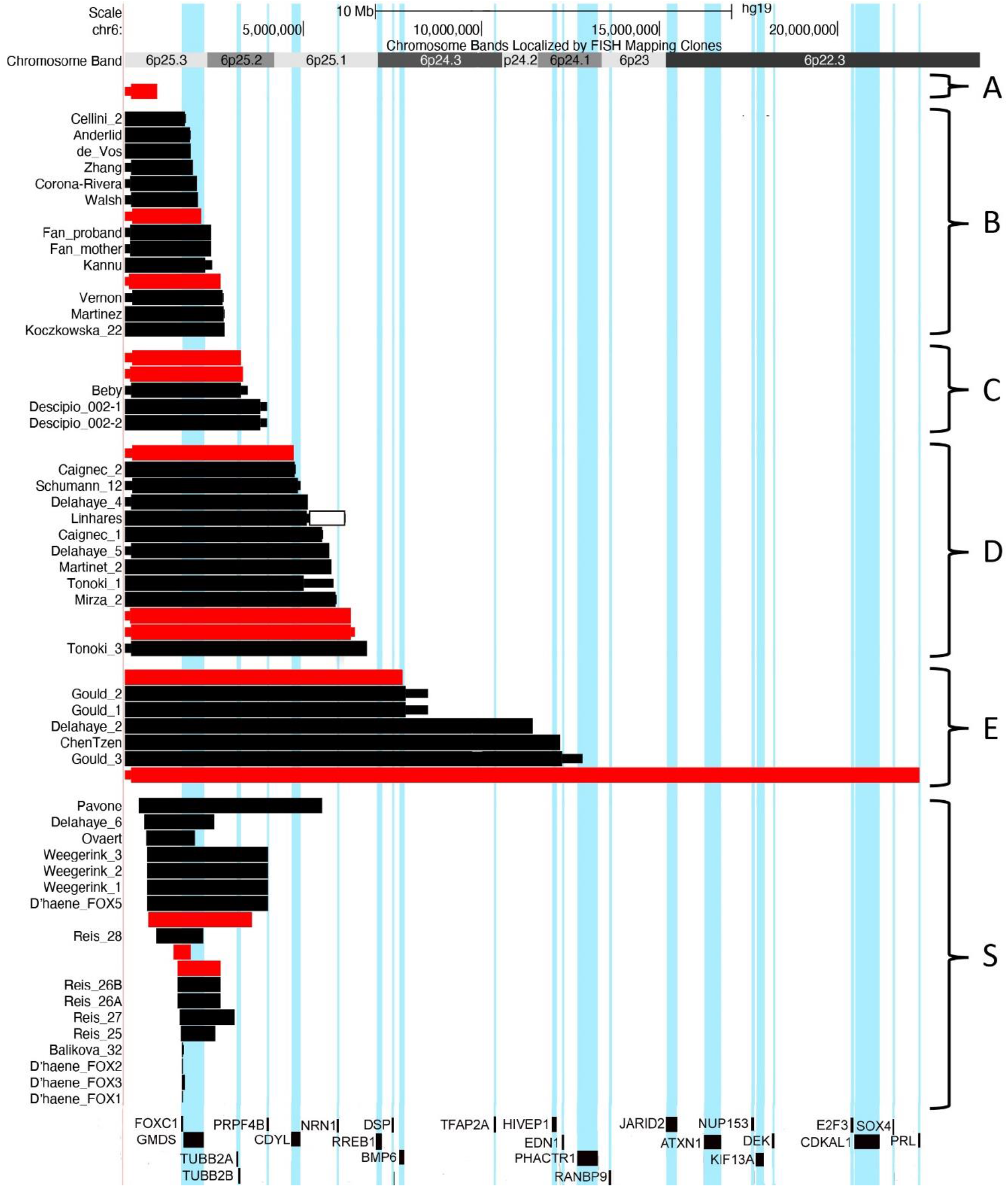
Overview of all terminal and subterminal 6p deletions. Deletions from our parent cohort (red bars) and literature cohort (black bars) are divided into six subgroups (A–S). A: terminal deletion not including *FOXC1* (breakpoint distal to 1.61 Mb), B: terminal deletions including *FOXC1* and not including *TUBB2A* (breakpoint between 1.61 and 3.15 Mb), C: terminal deletions including *TUBB2A* and not including *PRPF4B* (breakpoint between 3.15 and 4.02 Mb), D: terminal deletions including *PRPF4B* and not including *RREB1* (breakpoint between 4.02 and 7.11 Mb), E: terminal deletions including *RREB1* (breakpoint proximal to 7.11 Mb) and S: subterminal 6p deletions (distal breakpoint proximal to 0.35 Mb and including at least part of *FOXC1*). The literature cases were derived from 28 reports [2,4–8,21–42]. All deletions are visualised using the UCSC genome browser (GRCh37/hg19) (https://genome.ucsc.edu) [13]. Minimum and maximum breakpoints of deletions are visualised, when available. The literature case of Linhares et al. also has a duplication (indicated by a white bar).

### Participant and genotype characteristics

As of August 2022, the Chromosome 6 Questionnaire had been completed for 134 individuals. This included 13 individuals with a terminal (n = 10) or subterminal (n = 3) 6p deletion who could be included in our parent cohort. We further included 46 individuals from 28 published papers [2,4–8,21–42], 30 with a terminal and 16 with a subterminal deletion, who constitute our literature cohort. We thus included 59 patients in total: 40 with a terminal 6p deletion and 19 with a subterminal 6p25 deletion. Two literature cases [43,44] were excluded because their deletion was a benign variant according to the DGV, and one of these individuals appeared to have received a different genetic diagnosis since the original publication (personal communication Koolen et al.). Six of the included individuals exhibited small chromosomal rearrangements in addition to their terminal or subterminal 6p deletion, which we accepted based on size and gene content. Three terminal deletions (two literature cases [6,21] and one parent case (patient A) were the result of a ring chromosome 6. For details on the age and sex of all the individuals in our cohorts, see Table 1. The literature cohort included three fetuses [5,37,42] and three patients of unknown age [4,30] who were not included in the age calculations.

**Table 1.** Patient and genotype characteristics of 6p terminal and subterminal deletion subgroups

| Subgroup<sup>1</sup> | Cohort<br>Parent/<br>Literature | Sex<br>Female/<br>Male | Age in<br>years;<br>months<br>median<br>(range)<sup>2</sup> | Deletion<br>size in Mb<br>median<br>(range) | No. of<br>OMIM<br>genes<br>range<br>(median) | No. of HI-genes<sup>3</sup><br>range<br>(median) |
| --- | --- | --- | --- | --- | --- | --- |
| Patient A<br>(n = 1) | 1/0 |  |  |  | 4 | 0 |
| Terminal B<br>(n = 14) | 2/12 | 13/1 | 14;0 (n = 13)<br>(2;1–53;0) | 2.21<br>(1.68–2.8) | 9–10<br>(9) | 2 |
| Terminal C<br>(n = 5) | 2/3 | 2/3 | 2;3 (n = 5)<br>(0;10–20;0) | 3.31<br>(3.24–3.81) | 18–20<br>(20) | 4 |
| Terminal D<br>(n = 13) | 3/10 | 7/6 | 5;0 (n = 12)<br>(0;3–15;7) | 5.54<br>(4.74–6.79) | 24–31<br>(28) | 6–7 |
| Terminal E<br>(n = 7) | 2/5 | 4/2 | 2;0 (n = 3)<br>(0;1–2;4) | 11.45<br>(7.81–22.31) | 37–88<br>(55) | 10–24<br>(11) |
| Total terminal<br>(n = 40) | 10/30 | 26/13 | 5;8 (n = 34)<br>(0;1–53;0) | 4.28<br>(0.92–22.31) | 4–88<br>(22) | 0–24<br>(5) |
| Subterminal S<br>(n = 19) | 3/16 | 10/7 | 7;5 (n = 19)<br>(0;1–49;0) | 1.27<br>(0.005–5.13) | 1–27<br>(5) | 1–6<br>(2) |

<sup>1</sup> For more information on the division of subgroups, see Methods and Figure 1.
<sup>2</sup> If known, fetuses excluded.
<sup>3</sup> HI = haploinsufficiency.
HI-genes are defined as genes with a HI score of 0-10% or a pLI score $\geq 0.9$ .

## Results

A summary of the most commonly reported features can be found in Table 2, with a more detailed overview given in Table S3, Additional File 3. Clinical photographs can be found in Figure 2 (*figure removed*). The relationship between specific clinical features, deletion size and gene content was visualised using the UCSC genome browser (https://genome.ucsc.edu) [13] and is shown in Figures 3 and 4 and supplementary Figures S1-S5, Additional File 2.

**Table 2.** Most prominent characteristics seen in individuals with terminal and subterminal 6p deletions

| Phenotypic characteristic |  |  |  |  |  |  | Sub-<br>terminal (S) | Total terminal<br>and<br>subterminal |
| --- | --- | --- | --- | --- | --- | --- | --- | --- |
|  | A<br>(n=1) | B<br>(n=14) | C<br>(n=5) | D<br>(n=13) | E<br>(n=7) | Total<br>terminal<br>(n=40) | (n=19) | (n=59) |
| Neonatal asphyxia | + | 0/6 | 0/2 | 2/3 | 1/2 | 4/14 | 0/3 | 4/17 |
| Neonatal feeding problems | + | 2/6 | 1/2 | 3/3 | 2/2 | 9/14 | 3/3 | 12/17 |
| Stature: Short/normal/long | Short | 3/7/0 | 0/4/0 | 0/10/1 | 1/1/0 | 5/22/1 | 3/5/0 | 8/27/1 |
| Micro-/Normo-/Macrocephaly | Micro-<br>cephaly | 1/6/2 | 0/4/0 | 0/9/2 | 1/1/0 | 3/20/4 | 0/5/0 | 3/25/4 |
| Large fontanelles | - | 1/5 | 0/1 | 1/4 | 1/2 | 3/13 | 2/2 | 5/15 |
| **Eyes and vision** |  |  |  |  |  |  |  |  |
| Hypertelorism | - | 9/11 | 4/5 | 12/12 | 7/7 | 32/36 | 8/8 | 40/44 |
| Abnormalities of vision | + | 8/9 | 4/5 | 9/9 | 2/3 | 24/27 | 7/7 | 31/34 |
| Refraction problems | - | 2/4 | 3/3 | 6/7 | 1/2 | 12/17 | 4/5 | 16/22 |
| Eye movement abnormalities | - | 6/10 | 1/3 | 7/9 | 1/4 | 15/27 | 3/5 | 18/32 |
| Glaucoma | - | 4/13 | 1/3 | 1/11 | 1/7 | 7/35 | 12/19 | 19/54 |
| Coloboma | - | 0/13 | 1/3 | 1/11 | 1/7 | 3/35 | 2/19 | 5/54 |
| Anterior segment dysgenesis | - | 8/12 | 2/3 | 10/10 | 4/6 | 24/32 | 14/18 | 38/50 |
| Axenfeld / Rieger anomaly | n.a. | 8/8 | 2/2 | 9/10 | 3/4 | 22/24 | 14/14 | 36/38 |
| Corneal opacity | n.a. | 2/8 | 0/2 | 2/10 | 2/4 | 6/24 | 0/14 | 6/38 |
| **Ears, hearing and balance** |  |  |  |  |  |  |  |  |
| Dysplastic and/or low-set ears | - | 3/7 | 0/3 | 4/12 | 6/7 | 13/30 | 2/5 | 15/35 |
| Abn. middle and/or inner ear | - | 2/4 | 2/3 | 3/4 | 2/2 | 9/14 | 3/5 | 12/19 |
| Abnormal cochlea | n.a. | 1/2 | 1/2 | 0/3 | 1/2 | 3/9 | 2/3 | 5/12 |
| Abn. tympanic membrane | n.a. | 1/2 | 0/2 | 1/3 | 0/2 | 2/9 | 2/3 | 4/12 |
| Hearing impairment | - | 9/12 | 2/3 | 7/11 | 4/4 | 22/31 | 10/13 | 32/44 |
| Balance problems | + | 3/4 | 1/3 | 3/3 | 0/2 | 8/13 | 3/6 | 11/19 |
| **Oral region** |  |  |  |  |  |  |  |  |
| Cleft lip and/or palate | - | 0/7 | 0/3 | 3/10 | 2/5 | 5/26 | 1/2 | 6/28 |
| Dental problems | - | 4/9 | 2/3 | 5/8 | 0/0 | 11/21 | 4/9 | 15/30 |
| **Internal organs and immune system** |  |  |  |  |  |  |  |  |
| Feeding difficulties | - | 1/4 | 2/3 | 3/3 | 2/2 | 8/13 | 2/3 | 10/16 |
| Lower intestinal problems | - | 1/3 | 0/2 | 2/3 | 1/2 | 4/11 | 1/3 | 5/14 |
| Congenital heart defect | + | 3/11 | 2/3 | 6/11 | 4/5 | 16/31 | 6/11 | 22/42 |
| Septal defect/PFO | + | 2/3 | 1/2 | 5/6 | 4/4 | 13/16 | 4/6 | 17/22 |
| Patent ductus arteriosus | - | 1/3 | 0/2 | 1/6 | 1/4 | 3/16 | 0/6 | 3/22 |
| Abnormal heart valves | - | 1/3 | 2/2 | 1/6 | 1/4 | 5/16 | 4/6 | 9/22 |
| Kidney abnormalities | - | 1/3 | 0/2 | 1/3 | 3/4 | 5/13 | 1/5 | 6/18 |
| Micropenis | - | 1/1 | 0/2 | 1/5 | 1/2 | 3/11 | 1/3 | 4/14 |
| Recurrent infections | + | 3/3 | 2/3 | 2/4 | 0/2 | 8/13 | 4/4 | 12/17 |
| **Skeletal abnormalities** |  |  |  |  |  |  |  |  |
| Scoliosis | n.a. | 2/4 | 0/3 | 1/4 | 2/4 | 5/16 | 1/3 | 6/19 |
| Hip dysplasia | u | 0/3 | 0/2 | 2/5 | 0/1 | 2/11 | 3/4 | 5/15 |
| Joint hypermobility | - | 2/3 | 0/2 | 3/4 | 1/3 | 6/13 | 4/4 | 10/17 |
| Pes planus/positional foot deformity | + | 2/10 | 1/4 | 4/8 | 3/6 | 11/29 | 2/6 | 13/35 |
| Bone deformities (epiphyseal) | u | 5/5 | 0/1 | 5/5 | 1/1 | 11/12 | 2/4 | 13/16 |
| **Nervous system** |  |  |  |  |  |  |  |  |
| Brain abnormalities on imaging | + | 7/8 | 4/4 | 9/11 | 7/7 | 28/31 | 8/9 | 36/40 |
| Cerebellar abnormality | - | 2/7 | 2/4 | 4/9 | 7/7 | 15/28 | 1/8 | 16/36 |
| Dandy-Walker complex | n.a. | 0/1 | 2/2 | 2/4 | 5/5 | 9/12 | 1/1 | 10/13 |
| Abnormal corpus callosum | - | 0/7 | 0/4 | 3/9 | 4/7 | 7/28 | 2/8 | 9/36 |
| Ventriculomegaly/hydrocephalus | - | 3/8 | 3/4 | 5/9 | 5/7 | 16/29 | 3/8 | 19/37 |
| Abnormal white matter | u | 5/6 | 0/4 | 5/9 | 0/5 | 10/24 | 6/7 | 16/31 |
| Seizures | - | 1/4 | 1/2 | 2/4 | 1/3 | 5/14 | 1/2 | 6/16 |
| Hypotonia | + | 3/3 | 4/4 | 7/9 | 2/2 | 17/19 | 3/5 | 20/24 |
| Sensory impairment | + | 0/1 | 1/2 | 2/4 | 1/2 | 5/10 | 0/2 | 5/12 |
| **Development and behaviour** |  |  |  |  |  |  |  |  |
| Developmental delay | - | 11/14 | 2/3 | 10/10 | 1/1 | 24/29 | 10/12 | 34/41 |
| Sleeping problems | + | 0/2 | 0/2 | 2/3 | 0/2 | 3/10 | 2/2 | 5/12 |
| Social behaviour | + | 2/4 | 2/3 | 2/6 | 0/2 | 7/16 | 2/4 | 9/20 |
| Quiet behaviour | - | 0/4 | 1/3 | 3/6 | 1/2 | 5/16 | 1/4 | 6/20 |
| Autistic behaviour/spectrum | - | 1/2 | 2/3 | 1/4 | 0/2 | 4/12 | 2/4 | 6/16 |

**Fig. 3.**
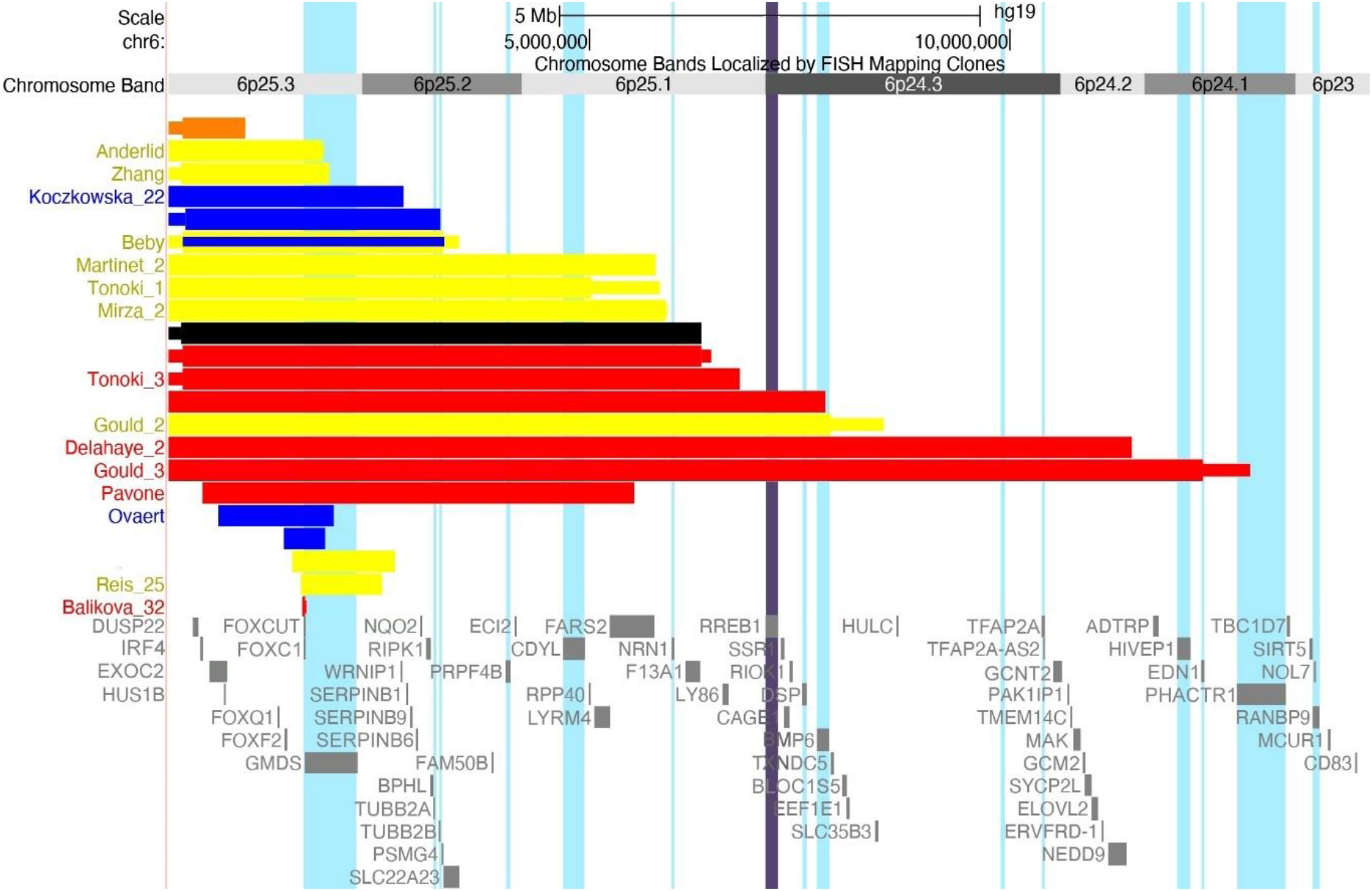
Overview of patients presenting with heart defects. Ventricular septal defect (orange), atrial septal defect or patent foramen ovale (yellow), cardiac valve abnormality (blue), complex heart defect, including septal defects and valve abnormalities, or coarcation of aorta, or hypoplastic left heart (red) and type unknown (black). The gene *RREB1* (see Discussion) is indicated by a purple vertical bar.

**Fig. 4.**
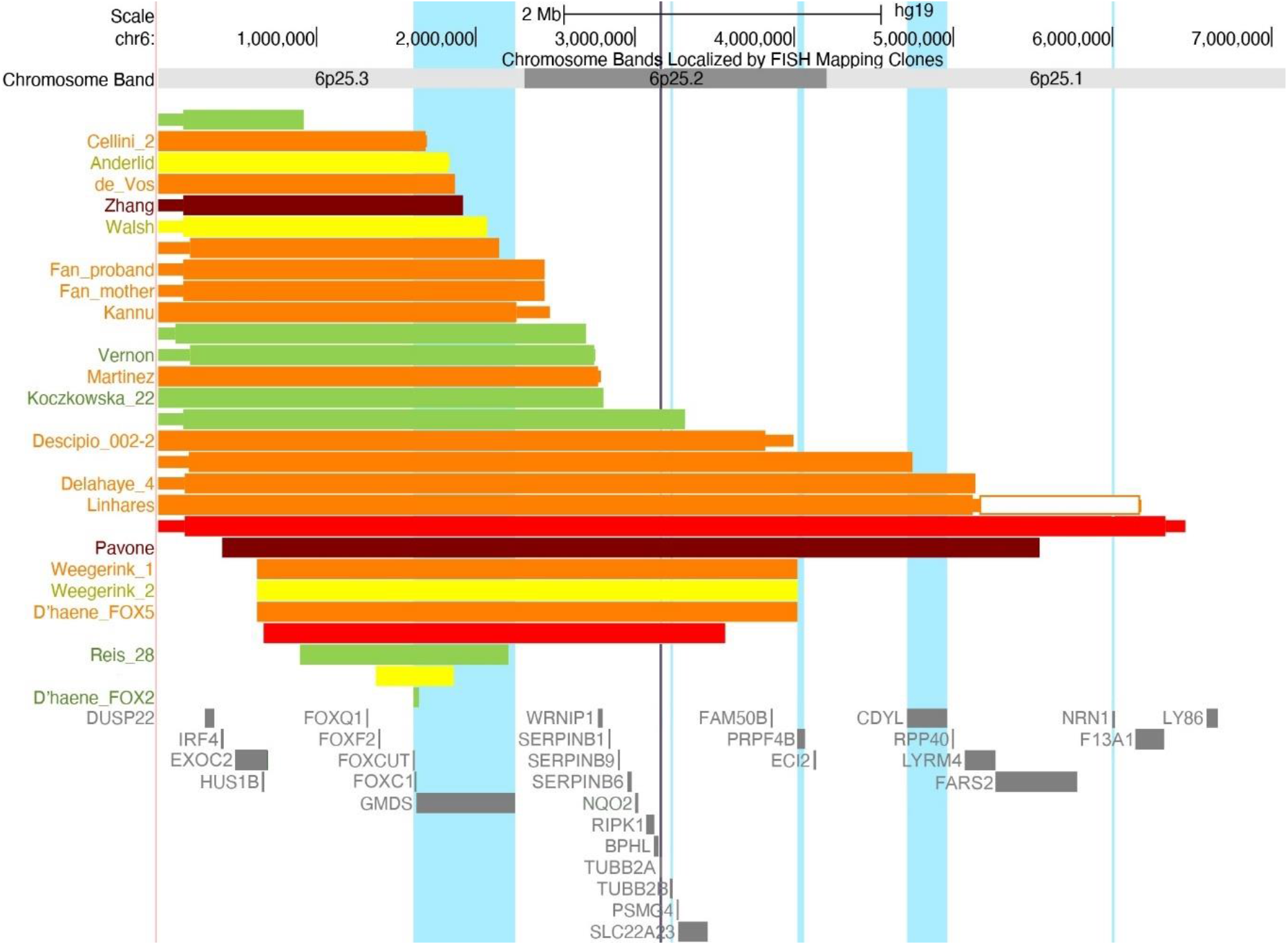
Overview of individuals for whom development could be categorized. They were categorized as having normal development (IQ > 85, green), borderline (IQ 70–85, yellow), mild (IQ 50–70, orange), moderate (IQ 30– 50, red) or severe (IQ < 30, dark red) delay. The gene TUBB2A is indicated by a purple vertical bar (see Discussion).

### Differences between cohorts

The availability of phenotype data for the two cohorts is also presented in Table S3, Additional File 3. Phenotype data that was present for almost all individuals in the parent cohort but only present for less than 25% of the literature cohort included, amongst others, neonatal problems, large fontanelles, middle and/or inner ear anomalies, specific skeletal problems (such as joint hypermobility, hip dysplasia and abnormalities of the spinal column), gastrointestinal and kidney problems, recurrent infections, seizures, sleeping problems, developmental milestones and behavioural characteristics. In contrast, information that was available to a lesser extent in the parent cohort, but still for more than 25% of the cases, was mostly on very specific features such as the type of cerebellar abnormality and the presence of white matter abnormalities. For data that was sufficiently present for both cohorts (available for at least 25% of the individuals in each cohort), the clinically relevant differences concerned the severity of vision problems and hearing impairment and the presence of eye movement abnormalities and their subtypes (see below).

### Neonatal period, growth and facial characteristics

Neonatal feeding problems were reported in 12/17 individuals (10/13 from the parent cohort) and were more common in the larger (D–E) and subterminal (S) deletions (8/8 in D–E and S vs 4/9 in A– C). Neonatal asphyxia (4/17) was reported in subgroups A (1/1) and D–E (3/5), exclusively in the parent cohort.

Most participants were of normal height (27/36), but eight showed short stature, which was mostly seen in subgroups B (3/10) and S (3/8). Head circumference was normal in most individuals (25/32), but microcephaly (3/27) and macrocephaly (4/27) were also seen in terminal deletions (subgroups A–E). Macrocephaly was accompanied by ventriculomegaly and/or hydrocephalus in 2/3 individuals. Notably, large fontanels were reported in 5/15 participants (4/10 in the parent cohort) and could not be related to head size.

Commonly observed facial characteristics in all subgroups were hypertelorism (40/44; 91%, 68–93%), dysplastic outer or low-set ears (15/35) and dental problems (15/30). A cleft lip and/or palate was reported in five individuals with a larger terminal deletion (D–E) (Figure S1, Additional File 2) and in just one individual with a subterminal deletion.

### Eyes and vision

Eye abnormalities were present in subgroups B–S, with detailed information available for both cohorts (see Table S3, Additional File 3). Morphological eye abnormalities were mostly within the spectrum of anterior segment dysgenesis (ASD) (38/50; 76%, 64–80%) and included Axenfeld and/or Rieger anomaly (36/38; 95%) and corneal opacity (6/38; 16%), with corneal opacity exclusively seen in terminal deletions. Glaucoma was present in 19/54 (35%, 32–41%) and was more common in subgroup S (12/19). Vision problems (31/34) were mostly mild (18/31) and included predominantly abnormalities of refraction (16/22; 73%, 52–81%). When present, visual impairment was described more often as severe in the parent cohort (4/11) compared to the literature cohort (1/20). Eye movement abnormalities (18/32) were seen more commonly in literature cases and included strabismus (11/18; 61%), which was exclusively reported in literature, and nystagmus (5/18; 28%).

### Ears, hearing and balance

Abnormalities of the middle and/or inner ear were reported in 12/19 individuals and mainly included abnormalities of the cochlea (5/12) and the tympanic membrane (4/12). These were always combined with hearing impairment (32/44; 73%, 54–80%), but were not present in four patients who did not have hearing loss (parent cohort) (Figure S2, Additional File 2). Hearing impairment occurred in all subgroups and was mostly mild to moderate (12/18). Severe to profound hearing loss (6/18) was exclusively seen in individuals in the literature cohort, only one of whom was known to have middle and/or inner ear problems. Balance problems were seen in 11/19 individuals.

### Problems related to the internal organs

Feeding difficulties (10/16) were more common in subgroups C–E and S, and tube feeding was more often required in these subgroups. Gastrointestinal reflux (4/13) and lower intestinal problems (5/14) were also reported, almost exclusively in the parent cohort (Table S3, Additional File 3).

Congenital heart defects (22/42; 52%, 37–66%) mostly included abnormalities of the cardiac septa (17/22; 77%) and/or valves (9/22; 41%) (Table 2, details in Table S3 and Figure 3). Remarkably, complex heart defects were only seen in individuals with a deletion extending beyond position 5.54 Mb (subgroups D, E and S). Isolated heart valve abnormalities were seen in deletions smaller than 3.31 Mb (subgroups A–C), while septal defects occurred in all subgroups.

Kidney abnormalities (6/18) were particularly common in individuals of subgroup E (3/4). Recurrent infections were observed in 12/17 patients and mostly included ear (10/12; 83%) and/or respiratory tract (7/12; 58%) infections, when this information was available.

### Skeletal problems

Joint hypermobility (10/17) and positional foot deformities (13/35) were among the most common skeletal problems, with no notable differences between subgroups. In 4/5 individuals with pes planus and 2/8 individuals with other positional foot deformities, hypotonia was known to be present (Figure S3, Additional File 2). Bone deformities, including tubular bone (epiphyseal) dysplasia and delayed bone ossification, were present in 13/16 individuals and consisted of abnormalities of the femur, ulna and humerus (Table 2 and Table S3, Additional File 3). Scoliosis was reported in 6/19 individuals (exclusively in the literature cohort), and hip dysplasia was reported in 5/15 individuals.

### Neurological problems

Brain imaging was performed in 40 individuals using different techniques: MRI in 26, CT in 5, brain ultrasound in 3 and using an unknown imaging modality in 6 literature cases. The abnormalities reported (36/40; 90%, 61–93%) mostly included ventriculomegaly/hydrocephalus (19/37; 51%), cerebellar anomalies (16/36; 44%) and white matter abnormalities (16/31; 52%, 52–68%). The most commonly observed cerebellar anomalies were part of the Dandy-Walker complex (10/13; 77%), including hypoplasia of the cerebellar vermis (Dandy-Walker variant) and its combination with a dilated fourth ventricle and enlarged posterior fossa (Dandy-Walker malformation). Remarkably, a Dandy-Walker malformation/variant was only seen in deletions extending proximally beyond position 3.27 Mb (subgroups C–E and S), and its penetrance seemed higher in larger deletions (Figure S4, Additional File 2). Of the 19 patients with ventriculomegaly/hydrocephalus, 11 also had a Dandy-Walker complex anomaly (9) or an unspecified cerebellum malformation (2). White matter abnormalities did not seem to be related to deletion size. Corpus callosum abnormalities (9/36; 25%) were also seen, mostly in individuals with larger deletions (subgroups D–E) (Figure S5, Additional File 2).

Frequently observed neurological findings included hypotonia (20/24), sensory impairment (5/12) and seizures (6/16), which occurred in all subgroups. Abnormal pain sensation was reported by parents in subgroups D (3/3) and S (1/2).

### Development and behaviour

Developmental delay (34/41; 83%, 58–88%) was mostly mild. Table 3 provides an overview of the extent of developmental delay (if known), and this is also visualised for each deletion in Figure 4. There is no obvious correlation between the size of the deletion and the severity of the developmental delay, although normal development and borderline delay are only observed in individuals with a terminal deletion smaller than 3.31 Mb (subgroups A–C). The ages of achievement for some specific milestones are summarised in Table 4 and visualised in Figure 5. All children for whom this information was available could walk independently from the age of 4 years and could use two-word sentences by the age of 6. There were no obvious differences between subgroups in the ages at which these milestones were achieved.

**Table 3.** Development in patients with terminal and subterminal 6p deletions

|  | A | B | C | D | E | Total<br>terminal | Subterminal |
| --- | --- | --- | --- | --- | --- | --- | --- |
| Normal | 1 | 3 | 1 | 0 | 0 | 5 | 2 |
| Borderline |  | 2 | 0 | 0 | 0 | 2 | 2 |
| Mild delay |  | 7 | 1 | 3 | 0 | 11 | 2 |
| Moderate delay |  | 0 | 0 | 1 | 0 | 1 | 1 |
| Severe delay |  | 1 | 0 | 0 | 0 | 1 | 1 |
| Delayed but not specified |  | 1 | 1 | 6 | 1 | 9 | 4 |
| Unknown due to young age<sup>1</sup> |  | 0 | 1 | 3 | 3 | 7 | 4 |

Development is visualised per individual in Figure 4.
<sup>1</sup> Younger than 2 years of age.

**Table 4.** Age at achievement of specific milestones

| Milestone | Number of Individuals<sup>1</sup> | Median age of achievement in months (range) |
| --- | --- | --- |
| Sitting-up unaided | 7 | 11 (10–18) |
| Walking unsupported | 17 | 22 (16–42) |
| Building tower of 6 blocks | 6 | 26 (11–60) |
| Speaking first words | 9 | 18 (9–42) |
| Speaking two-word sentences | 6 | 30 (18–48) |

<sup>1</sup>For calculation of median ages and ranges, we only included individuals for whom the exact ages of achievement of the milestones were known.

**Fig. 5.**
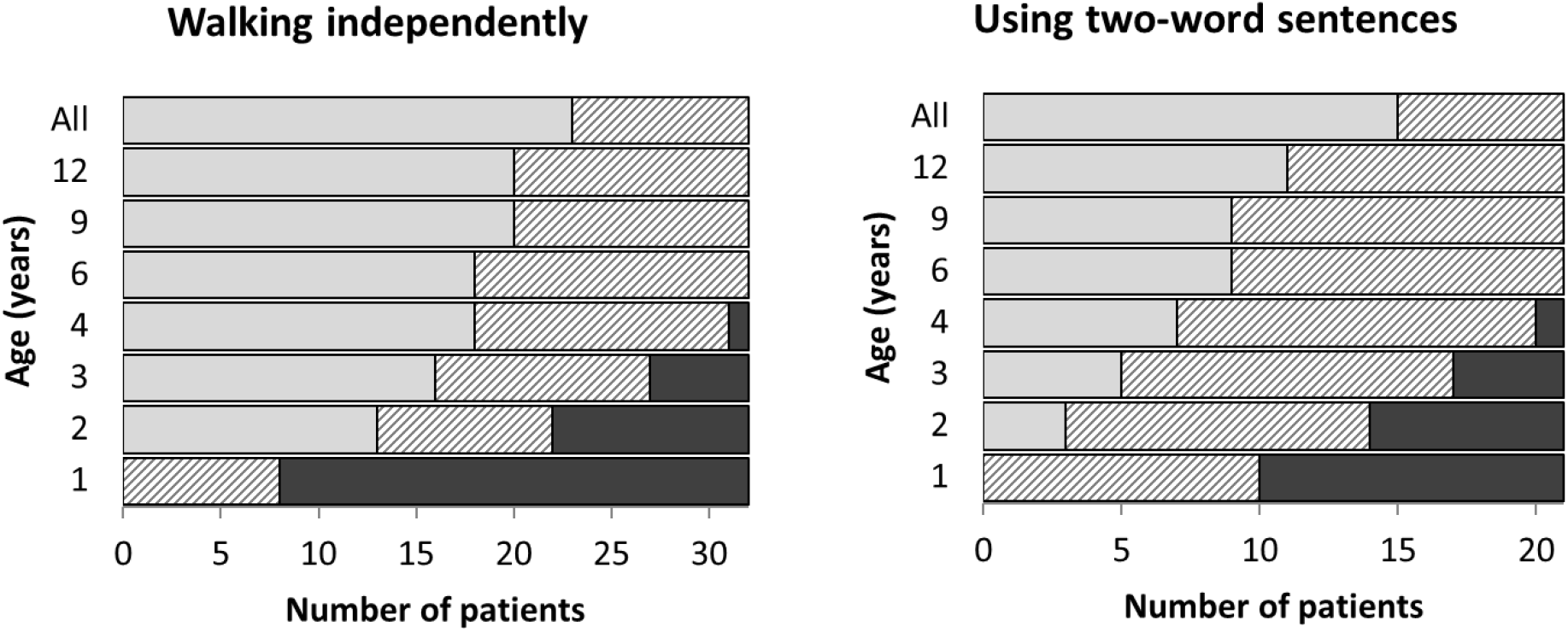
Age of achievement of milestones ‘walking independently and ‘using two-word sentences’ for the total group. Dark grey bars represent the number of children (x axis) that were not able to perform these milestones at that age (y axis). Light grey bars display the number of children who could perform these milestones at that age. Hatched bars represent children who could not perform the milestones but had not yet reached the age displayed on the y axis and children who are able to perform the milestones but for whom it is not known whether they had achieved them at the age on the y axis. In the top bar “All”, the light grey bar represents all the children who achieved the milestone, including those for whom the age of achievement is not known and the hatched bar represents children who have not yet achieved the milestone (but these children were all 4 years or younger).

Five parents reported that their child had sleep disturbances (5/12), mostly insomnia (5/5). Behaviour was most often described as social (9/20) and quiet (6/20), with aggressive behaviour seen in 3/20 individuals. Behavioural features within the autism spectrum were also reported in 6/16.

## Discussion

We analysed the clinical features reported for 59 individuals with a terminal or subterminal 6p25 deletion with the aim of providing physicians and parents with guidelines for clinical surveillance. In this discussion, we summarise the most common phenotypic features, elaborate on their clinical implications and provide recommendations. We also discuss how some of the phenotypic differences between subgroups may be explained by the deletion of specific genes in the 6pter region. Lastly, but importantly, we highlight the advantages of collecting phenotype data directly from parents.

### Phenotypes of terminal and subterminal 6p deletions

Individuals with terminal and subterminal 6p deletions including *FOXC1* appear to have a common phenotype, mainly consisting of ocular ASD, mild vision problems, brain malformations, congenital defects of the cardiac septa and valves, mild to moderate hearing impairment, eye movement abnormalities, hypotonia, mild developmental delay and dysmorphic features. Brain malformations included abnormalities of the cerebellum as part of the Dandy-Walker complex, ventriculomegaly and abnormalities of the white matter and corpus callosum. The most commonly reported dysmorphic features were hypertelorism, dental problems and outer ear anomalies. Other features of high clinical significance included skeletal abnormalities, middle and/or inner ear problems, recurrent infections and, to a lesser extent, seizures and glaucoma (often as a complication of the ASD). The parent cohort reported a number of new features that were only rarely mentioned in the literature as either present or absent. These included gastrointestinal problems, an abnormal pain sensation, balance problems and sleep disturbances. Behavioural characteristics were also primarily available for individuals in the parent cohort, who were most often described as being social and quiet.

Certain clinical features were more common in individuals with larger deletions. Feeding difficulties during and after the neonatal period were seen more often in deletions of subgroups C–E. Complex heart defects, corpus callosum abnormalities and cleft lip and/or palate were more commonly seen in subgroups D–E (deletion breakpoint proximal to 4.02 Mb). Kidney abnormalities were more common in individuals with the largest deletions (subgroup E), and the penetrance of cerebellar malformations within the Dandy-Walker spectrum also seemed higher in this subgroup. Developmental delay and its severity did not seem to correlate with deletion size, although penetrance seemed higher in individuals with deletions larger than 3.31 Mb (Figure 4).

No clinically relevant phenotypic differences were seen between terminal and subterminal deletions, apart from glaucoma, which was seen more often as a complication of Axenfeld-Rieger anomaly in the subterminal group (9/14) compared to the terminal group (5/24). Remarkably, corneal opacity was reported solely in the terminal deletions.

### The role of *FOXC1* in 6p25 deletion syndrome

6pter-p24 deletion syndrome, often referred to as 6p25 deletion syndrome, is a recognised clinical entity characterised by a spectrum of anterior chamber abnormalities combined with extra-ocular manifestations [2,3]. *FOXC1* is considered to be the most determinant gene for this syndrome, and interstitial 6p25 deletions that do not include *FOXC1* are considered to be different developmental disorders than terminal or subterminal 6p deletions that do encompass *FOXC1* [2]. *FOXC1* encodes a transcription factor that is active during the development of the eye, and pathogenic *FOXC1* variants are known to result in Axenfeld-Rieger syndrome (ARS) (MIM#602482). ARS is an autosomal dominant, genetically heterogeneous and clinically variable syndrome that is mainly characterised by developmental eye defects and specific extra-ocular features [45]. There is a high degree of phenotypic overlap between ARS and 6p25 deletion syndrome. The extra-ocular features seen in *FOXC1*-related ARS include many that are also present in 6p25 deletion syndrome, namely tooth abnormalities, sensorineural hearing loss, congenital heart defects, cerebellar abnormalities and craniofacial dysmorphisms such as maxillary hypoplasia and hypertelorism [2,45]. As seen from Table 2 and Table S3 (Additional File 3) eye abnormalities were generally well described in the included literature cases, and ocular ASD was one of the most common phenotypes seen in the total group. Next to the ocular phenotype, many individuals in our cohorts also show one or more of the other features of ARS. We therefore conclude that the penetrance of this syndrome in 6p25 deletions including *FOXC1* is high, albeit not complete.

Only one of our patients (patient A) had a terminal 6p deletion that did not involve *FOXC1*, and this patient did not have the Axenfeld-Rieger ocular phenotype (Figure 1). In literature, two patients have been described with eye abnormalities and a terminal 6p deletion that did not include *FOXC1* [43,44]. It was suggested that their eye abnormalities could be caused by deletion of *DUSP22* or disrupted regulatory elements of *FOXC1*. However, deletions including only *DUSP22* are qualified as benign in the DGV. We contacted Koolen et al. [44] about his patient and learned that, after publication, the patient was re-evaluated by SNP array, which confirmed that the 6p25 variant was benign and identified a pathogenic deletion of *EHMT1* (euchromatic histone methyltransferase 1 MIM*607001, chr 9q34.3), leading to a diagnosis of Kleefstra syndrome (MIM#610253) (personal communication Koolen et al.). This leads to speculate that there is also another (molecular) cause for the reported phenotype in the patient described by Chen et al. [43] and that haploinsufficiency of *FOXC1* remains the most likely cause for the ocular phenotype seen in 6p25 deletions.

### Extra-ocular features in the 6p25 deletion syndrome

Numerous extra-ocular features associated with 6p25 deletion syndrome have previously been related to haploinsufficiency of *FOXC1* [1–3,5]. We will therefore focus our discussion on how our findings support this connection, but we will also reflect on possible roles for haploinsufficiency of other genes in explaining the clinical differences between our subgroups. All but one individual in our cohort shared a deletion of *FOXC1* and its neighbouring gene *GMDS* (GDP-mannose 4,6-dehydratase, MIM*602884). *GMDS* is a predicted HI-gene (HI score 3.84% and pLI score 0.99), but little is known about its role and no phenotypes of pathogenic variants have been reported in humans thus far. Therefore, we will not discuss the role of this gene in 6p25 deletion syndrome further.

#### Hearing impairment and middle/inner ear abnormalities

Hearing impairment was seen in all subgroups and ranged from none to profound. Hearing loss is one of the most common non-ocular features of *FOXC1*-related ARS, whereas it is not commonly observed in ARS of other genetic origins. *FOXC1* is highly expressed in the auditory and vestibular sensory epithelium in mammalian inner ears, and its haploinsufficiency might lead to hearing loss [7,46,47]. *FOXC1* was the only shared relevant HI-gene in the individuals with hearing impairment in our cohorts, and the only participant without a *FOXC1* deletion did not exhibit hearing impairment (Figure S2, Additional File 2), supporting the hypothesis that haploinsufficiency of *FOXC1* is related to the development of hearing impairment in 6p25 deletions.

The severity of hearing impairment in our cohort could not be linked to the size of the deletion (Figure S2, Additional File 2). The variability in severity might have a multifactorial cause, including factors such as recurrent otitis media and middle/inner ear abnormalities. Information on the presence of middle and/or inner ear abnormalities was scarce in the literature cohort, resulting in low numbers that prevented us from drawing any conclusions regarding its relation to hearing loss severity. All individuals with inner ear abnormalities in our cohorts missed one copy of *FOXF2* (forkhead box f2, MIM*603250), which has also been shown to play a role in ear development [48,49]. Hearing impairment was seen in all these individuals and ranged from mild to profound (when this information was available). None of the individuals whose deletion did not include *FOXF2* were reported to have inner ear abnormalities. Thus far, no heterozygous *FOXF2* variants with hearing impairment have been described in literature, and *FOXF2* is not a predicted HI-gene (HI score: 29.63%, pLI score 0.85). However, we cannot rule out that haploinsufficiency of *FOXF2* plays a role in the development of inner ear abnormalities and hearing loss in 6p25 deletion patients.

#### Skeletal defects

Multiple skeletal problems were observed in our patients, including an abnormal curvature of the vertebral column, joint hypermobility, hip dysplasia, positional foot deformities and epiphyseal dysplasia of the long tubular bones. These abnormalities were seen in all subgroups with a *FOXC1* deletion, irrespective of deletion size. Skeletal defects have often been described in combination with ARS [2,25,38], and it has been suggested that haploinsufficiency of *FOXC1* might play a role in their development [38], a theory also supported by studies in mice [50]. *FOXC1* is also required for calvarial bone development [51], and large fontanelles were observed in several of our participants. Our findings thus support the idea that the skeletal features seen in 6p25 deletion syndrome are also related to haploinsufficiency of *FOXC1*.

#### Cleft lip/palate

Orofacial clefting was seen in six patients. Of these, three had an isolated cleft palate and three also had a cleft lip (Figure S1, Additional File 2), which suggests a different embryological origin. The only relevant HI-gene shared among the individuals with an isolated cleft palate was *FOXC1*, which has not specifically been related to this phenotypic feature in the literature, and a cleft palate is not typically seen in *FOXC1*-related ARS. Another deleted gene shared among these patients is *FOXF2*, although it is not an HI-gene. Heterozygous *FOXF2* variants in humans have been associated with cleft palate [52], which has also been seen in *Foxf2* -/- mice [53]. However, the majority of patients in our cohort with a deletion including *FOXF2* did not have a cleft palate, demonstrating reduced penetrance of this feature.

Notably, a cleft lip and palate was seen only in individuals with terminal deletions exceeding 5.9 Mb (in subgroups D–E). Orofacial clefting has often been described in terminal 6p deletions with proximal breakpoints beyond the 6p25 region, but is rarely seen in smaller deletions [1,4,5,25,37], which suggests that a more proximally located gene might contribute to the development of this feature. The gene *OFCC1* (ofc1 candidate gene 1, MIM*614287), located at position 9,707,990– 9,939,582 and shared by two of the three individuals with a cleft lip and palate in our cohort, has been proposed to be possibly related to this feature [54]. Our findings support the idea that the contribution of a more proximally located gene to orofacial clefting might explain its higher prevalence in individuals with larger terminal 6p deletions.

#### Heart defects

*FOXC1* variants are often accompanied by heart defects [55,56] and could explain the heart defects seen in our cohorts (Figure 3). However, more complex heart defects were seen in some patients with larger deletions (subgroup E), including a hypoplastic left heart, atrial septal defect and ventricular defect in one fetus [5] and a coarctation of the aorta and a ventricular septal defect in another patient. The deletions in these patients also include the HI-gene *RREB1* (ras-responsive element binding protein 1 MIM*602209). Tassano et al. described a patient with a 6p25.1p24.3 deletion including *RREB1* who had a mild patent foramen ovale and dysplastic tricuspid valve [57]. Kent et al. reported a patient with a 6p25.1p24.3 deletion and associated the gene *RREB1* with Noonan syndrome, a syndrome in which both structural heart defects and cardiomyopathy are common. However, the Kent et al. patient only presented with a right bundle branch block, whereas the same study also showed that Rreb1 +/- mice developed cardiomyopathy within 6 months but had no structural heart defects [58]. Neither arrhythmia nor cardiomyopathy were seen in our cohorts. Within our institute, we previously identified a *RREB1* variant in a father and son presenting with a Noonan phenotype. Reanalysis of WES data in patients with congenital heart defects then revealed another six *RREB1* variants. These patients presented with a range of congenital heart defects, including transposition of the large arteries, coarctatio aortae, hypoplasia of the right ventricle, ventricular septal defect and/or valve abnormalities (own unpublished data). If these variants result in a loss-of-function effect with incomplete penetrance, it could explain the more complex heart defects in some of our patients with larger deletions that included both *FOXC1* and *RREB1*. However, patients with a *FOXC1* variant have also been reported with a tetralogy of Fallot or a coarctation [59,60]. For now, we conclude that the heart defects seen in 6p25 deletions may be related to haploinsufficiency of *FOXC1* but that there may also be an additive effect of *RREB1* in larger deletions.

#### Brain abnormalities and development

Several brain abnormalities were observed in all subgroups, confirming the role of *FOXC1* in the development of brain aberrations. Cerebellar abnormalities within the Dandy-Walker spectrum were among the most common brain abnormalities in our cohort, and they have been associated with *FOXC1* pathogenic variants in the literature [61]. It has been suggested that the size of the deletion correlates with the extent of the cerebellar malformation, with the classic Dandy-Walker malformation being more common in 6p25 deletions encompassing other genes besides *FOXC1* [2,61]. Interestingly, we observed cerebellar abnormalities within the Dandy-Walker spectrum for terminal deletions larger than 3.27 Mb (subgroups C–E) in our cohort. The deletions of all individuals with a Dandy-Walker malformation/variant also included the gene *TUBB2B* (tubulin, beta-2b MIM*612850) (Figure S4) known to cause tubulinopathies when haploinsufficient [62]. Pathogenic *TUBB2B* variants have been reported in patients with a range of cortical dysgeneses, with cerebellar vermis hypoplasia also seen in many of the reported cases [63,64]. We thus suggest that *TUBB2B* might play an additional role next to *FOXC1* in the development of Dandy-Walker spectrum abnormalities in 6p25 deletions. The severity within the Dandy-Walker spectrum in our cohorts could not be related to deletion size, but its penetrance seemed higher in the subgroup with the largest deletions (subgroup E) (Figure S4, Additional File 2).

Corpus callosum abnormalities were only seen in larger terminal deletions (subgroups D–E) and one patient with a subterminal deletion. Besides *FOXC1*, all individuals with this abnormality also had a deletion of the HI-genes *TUBB2A* (tubulin, beta-2a MIM*615101) and *TUBB2B*. Corpus callosum abnormalities have also been seen in patients with pathogenic variants in these genes, in combination with a range of other brain malformations [62,65]. A possible role for *TUBB2A* and *TUBB2B* in the cortical phenotype seen in 6p25 deletions was proposed previously [66], and we suggest that the corpus callosum abnormalities in our cohorts might be explained by the deletion of either or both of these genes (Figure S5, Additional File 2). The presence of the other brain abnormalities did not seem to relate to deletion size.

Developmental level was variable in the whole cohort (Table 3 and Figure 4). The only shared HI-gene previously associated with developmental delay is *TUBB2A*; individuals with pathogenic *TUBB2A* variants present with developmental delay [67]. In our cohort only 1/11 individuals with a deletion including *TUBB2A* had normal development, in contrast to 6/17 for patients without *TUBB2A* in their deletion (Figure 4). *TUBB2A* might thus explain the higher penetrance of developmental delay in patients with larger deletions. However, the cause of developmental delay in 6p25 deletions is most likely multifactorial, with brain abnormalities, hearing impairment and vision problems also playing a role.

### Recommendations for clinical surveillance and follow-up in terminal and subterminal 6p deletions

In Table 5, we present recommendations for the surveillance of patients with terminal and subterminal 6p deletions, which result in a common phenotype with variable penetrance. Our recommendations are for all individuals diagnosed with a terminal or subterminal 6p deletion including *FOXC1*, irrespective of deletion size unless otherwise stated.

**Table 5.** Clinical recommendations for surveillance in patients with terminal 6p deletions and subterminal 6p deletions including *FOXC1*

| Investigation | Upon diagnosis | Follow-up |
| --- | --- | --- |
| Thorough neurologic investigation | + | i |
| Brain MRI | + | i |
| EEG | i* | i* |
| Thorough ophthalmological investigation with special attention paid to the anterior segment | + | a |
| Refraction measurements | + | a |
| Eye-pressure measurement | + | a |
| Hearing assessment | + | a / i** |
| Imaging of the os petrosum | i*** | i*** |
| Cardiac ultrasound | + | i |
| Attention for lip and/or palate abnormalities | +, if deletion > 4.02 Mb | i |
| Attention for tooth abnormalities | + | a / i** |
| Referral to dental specialist | i | i |
| Attention for feeding problems and gastro-intestinal issues | + | + |
| Renal ultrasound | +, if deletion > 4.02 Mb | i |
| Be aware of recurrent ear and respiratory tract infections | + | + |
| Assessment of growth and posture | + | a / i** |
| Attention for foot deformities, scoliosis and epiphyseal dysplasia | + | + |
| Referral to orthopaedics and/or rehabilitation specialist | i | i |
| Neurodevelopmental assessment | + | i |
| Attention for sleeping problems | + | + |

i = upon indication, a = annually
\* In case of clinical suspicion of seizures or severe sleeping problems.
\*\*\* In case of hearing loss.

Upon diagnosis, a thorough neurological and ophthalmological investigation should be performed with special attention paid to the anterior segment of the eye, followed by annual follow-up of eye pressure and refraction. Glaucoma was seen in more than a third of the individuals in our cohort, and almost half of ARS patients develop glaucoma, so timely detection and intervention (if needed) are important in preventing permanent damage. A brain MRI and cardiac ultrasound should be carried out upon diagnosis and, if clinically indicated, upon follow-up. An EEG should be considered when seizures are clinically suspected or when there are severe sleeping problems. Hearing should be assessed at least annually at a young age and upon indication at later ages. If hearing loss is present, imaging of the os petrosum is recommended to determine optimal hearing restoration. If middle/inner ear abnormalities are present, a brain-anchored hearing aid (BAHA) may be a better-suited solution than behind-the-ear hearing devices. For terminal deletions exceeding 4.02 Mb, a kidney ultrasound should also be performed upon diagnosis, and extra attention should be given to the presence of lip and/or palate abnormalities, which may be missed if small or submucous.

Assessment of growth and posture should be performed annually, with special attention paid to positional foot deformities, scoliosis and an increased risk of epiphyseal dysplasia. Referral to an orthopaedic or rehabilitation specialist should be considered when skeletal abnormalities are present, as should referral to a dental specialist if tooth abnormalities are present. Other recommendations are less specifically related to (sub)terminal 6p deletions. These include attention to and treatment for feeding and gastrointestinal problems and recurrent infections, optimal support to optimise the child’s developmental abilities, attention to sleeping problems and appropriate support for child and parents if there are behavioural problems.

### Strengths and limitations

The phenotype associated with a chromosomal aberration is the result of a complex process and cannot simply be predicted based on the sum of the deleted genes. The continuous gain of knowledge on the haploinsufficiency effect of genes is a strong asset for linking chromosomal aberrations to certain clinical features, but as-of-yet unexplored factors such as position effects, the role of deleted non-coding parts of DNA that may include regulatory elements, or specific genes with yet unknown functions impede phenotypic characterisations based solely on gene content. The best way to gain a thorough understanding of the clinical consequences of deletions is thus to collect phenotype and genotype data on as many affected individuals as possible. In this study, we were able to do so by gathering data from a large cohort of affected individuals. We thus did not rely solely on information from literature reports; we also collected a considerable amount of data directly from families of newly identified patients, which has a number of advantages.

Firstly, as the Chromosome 6 Questionnaire covers a wide scope of characteristics, we were able to identify previously unrecognised clinical features, to describe the developmental aspects in more detail and to provide information on behavioural patterns seen in affected individuals, all of which are important information for parents. Secondly, collecting phenotype data through the Chromosome 6 Questionnaire helped us gain knowledge on clinical issues that are absent in patients with (sub)terminal 6p deletions, information that usually cannot be collected from literature. Knowledge of the absence of clinical features is essential in a clinical setting because it can prevent unnecessary investigations and provide reassurance to parents. Thirdly, collecting information directly from parents can result in a more realistic phenotype description because taking data only from literature reports can lead to an overestimation of the prevalence of certain features due to selection or publication bias. For instance, papers focusing on ARS may describe patients with a terminal 6p/*FOXC1* deletion who were identified because they had the ocular phenotype, resulting in an overestimation of this phenotype. Another advantage of collecting data directly from parents is that information can be easily updated or supplemented, as shown by the additional questionnaire we offered to our parent cohort, whereas case reports only provide information up to the point of publication. All in all, collaborating with patients and their families increases data availability [10,68] while also making a crucial step towards patient empowerment.

A well-known debate is whether information collected directly from families is accurate enough for scientific research or the development of evidence-based clinical guidelines. In our data consistency study, we compared the data collected from parents via the Chromosome 6 Questionnaire to data extracted from the medical files on the same individuals [69]. This revealed that data from these two sources was highly consistent [69]. In the current study, we did not find any major discrepancies in the phenotypic features between the parent and literature cohorts, with only the following exceptions: vision problems were more often described as severe by parents, while severe/profound hearing loss and eye movement abnormalities were more often described in the literature cases. However, it should be noted that we could only assess differences between the two cohorts for half of the observed features due to lack of data on numerous features in the literature cohort.

In our results, we presented the mean percentages for the prevalence of certain features, including a range with the minimum and maximum value. The mean percentage given here might be an overestimation of the prevalence of these characteristics because the presence of features that were not mentioned in the literature as either present or absent was considered unknown. However, it can be argued that many features, especially those of high clinical significance, are not present when not reported in the literature. For this reason, the minimum value of the range might be more realistic. Ultimately, to gain a better understanding of the prevalence of the clinical features seen in these aberrations and to help guide (future) parents, detailed clinical information must be collected from a greater number of new patients.

## Conclusions

We present a comprehensive overview of the clinical characteristics seen in terminal and subterminal 6p deletions, which appear to be largely attributable to haploinsufficiency of *FOXC1*. Penetrance, however, is variable, and not all features of ARS are present in all individuals. Furthermore, certain phenotypic features, namely complex heart defects, corpus callosum abnormalities, orofacial clefting and kidney abnormalities, were more commonly seen in individuals with deletions exceeding 4.02 Mb. Some of these differences might be explained by the haploinsufficiency of other genes in the terminal 6p region, such as *RREB1* in the cardiac phenotypes and *TUBB2A* and *TUBB2B* in the cerebral phenotypes. Furthermore, we have demonstrated the advantage of gathering clinical information directly from families, which enabled us to delineate the developmental abilities and identify previously unrecognised clinical features. Based on our observations in this large study group, we have provided clinical surveillance recommendations to aid healthcare professionals and patients and their families.

## Data Availability

The data presented in this study are openly available in DECIPHER at https://decipher.sanger.ac.uk (reference number IDs removed).

## List of abbreviations

ARS: Axenfeld-Rieger syndrome
ASD: Anterior segment dysgenesis
BAHA: Brain-anchored hearing aid
DGV: Database of Genomic Variants
HI: Haploinsufficiency
IQ: Intelligence quotient
pLI: probability of loss-of-function intolerance

## Declarations

### Ethics approval and consent to participate

The accredited Medical Ethics Review Committee of the University Medical Centre Groningen waived full ethical evaluation because, according to Dutch guidelines, no ethical approval is necessary if medical information that was already available is used anonymously and no extra tests have to be performed. Consent for participation was obtained from all patients involved in the study as part of the sign-up process for the Chromosome 6 Project.

### Consent for publication

Consent for the anonymous publication of medical information was obtained as part of the sign-up process for the Chromosome 6 Project. Consent for the publication of photographs was obtained separately.

### Availability of data and materials

The data presented in this study are openly available in DECIPHER at https://decipher.sanger.ac.uk, (*reference number IDs removed*).

### Authors’ contributions

Conceptualisation: CMARA and AE; methodology, MAS, CMARA and AE; software: MAS; validation: CMARA and AE; formal analysis, ER, WSKF and AE; resources, CMARA; data curation, TD and AE; writing—original draft preparation, ER; writing—review and editing, WSKF, MAS, TD, CMARA and AE; visualisation, ER and AE; supervision CMARA and AE; funding acquisition, ER, MAS, CMARA and AE. All authors have read and agreed to the published version of the manuscript.

## Acknowledgements

We would like to express our gratitude to all the patients and their families who participated in this study. We thank Pauline Bouman for being our contact for the Chromosome 6 Facebook group. We would like to thank our graduate students, Jennifer Geurink and Laura Monsma, for their contribution. We also thank Ni-Chung Lee and Elena Semina for providing details on the array results of their previously published patients, David Koolen for personal communication on his previously published patient, Kate McIntyre for editing the manuscript and the members of the MOLGENIS team at the Groningen Coordination Center, in particular Fernanda de Andrade and Mariska Slofstra, for their support on the online Chromosome 6 Questionnaire.

## Additional files

**Additional File 1.xlsx**

Table S1. (*removed*)

**Additional File 2.pdf**

Table S2. HI and pLI scores

Figure S1. Cleft lip and/or palate.

Figure S2. Hearing impairment and middle/inner ear abnormalities.

Figure S3. Positional foot deformity and pes planus.

Figure S4. Cerebellar abnormality and Dandy-Walker malformation.

Figure S5. Corpus callosum abnormality.

**Additional File 3.xlsx**

Table S3. Detailed overview of the phenotypic characteristics seen in individuals with (sub)terminal 6p deletions

