## Additional File 2 for "The phenotypic spectrum of terminal and subterminal 6p deletions based on a social media-derived cohort and literature review"

**Content**

**Tables:**

Table S2. HI and pLI scores

**Figures:**

Figure S1. Cleft lip and/or palate.
Figure S2. Hearing impairment and middle/inner ear abnormalities.
Figure S3. Positional foot deformity and pes planus.
Figure S4. Cerebellar abnormality and Dandy-Walker malformation.
Figure S5. Corpus callosum abnormality.

**Table S2.** HI and pLI scores

| Location | Gene | MIM* | % HI | pLI |
|---|---|---|---|---|
| 6p25.3 | DUSP22 | 616778 | 38.67 | 0.01 |
| | IRF4 | 601900 | 19.26 | 0.86 |
| | EXOC2 | 615329 | 34.25 | 0.00 |
| | HUS1B | 609713 | 97.37 | |
| | FOXQ1 | 612788 | 74.58 | 0.62 |
| | FOXF2 | 603250 | 29.63 | 0.85 |
| | **FOXC1** | 601090 | 9.00 | 0.95 |
| | **GMDS** | 602884 | 3.84 | 0.99 |
| 6p25.2 | WRNIP1 | 608196 | 36.94 | 0.02 |
| | SERPINB9 | 601799 | 88.03 | 0.01 |
| | SERPINB6 | 173321 | 69.00 | 0.0 |
| | NQO2 | 160998 | 69.71 | 0.00 |
| | RIPK1 | 603453 | 52.23 | 0.01 |
| | BPHL | 603156 | 69.91 | 0.00 |
| | **TUBB2A** | 615101 | 20.25 | 0.93 |
| | **TUBB2B** | 612850 | 24.97 | 0.99 |
| | PSMG4 | 617550 | 70.83 | 0.00 |
| | SLC22A23 | 611697 | 50.76 | 0.83 |
| | FAM60B | 614686 | 73.35 | 0.15 |
| | **PRPF4B** | 602338 | 3.37 | 1.00 |
| | ECI2 | 608024 | 64.53 | 0.00 |
| 6p25.1 | **CDYL** | 603778 | 42.51 | 1.00 |
| | RPP40 | 606117 | 51.93 | 0.00 |
| | PPP1R3G | 619541 | 87.24 | 0.00 |
| | LYRM4 | 613311 | 40.98 | 0.20 |
| | FARS2 | 611592 | 44.69 | 0.0 |
| | **NRN1** | 607409 | 7.67 | 0.86 |
| | F13A1 | 134570 | 36.60 | 0.0 |
| | LY86 | 605241 | 78.98 | 0.00 |
| 6p24.3 | **RREB1** | 602209 | 46.41 | 1.00 |
| | SSR1 | 600868 | 28.18 | 0.27 |
| | CAGE1 | 608304 | 84.08 | 0.00 |
| | RIOK1 | 617753 | 55.48 | 0.00 |
| | **DSP** | 125647 | 12.86 | 1.00 |
| | **BMP6** | 112266 | 3.67 | 0.76 |

All OMIM genes studied, extending from 6p25.3 to 6p24.3. Predicted HI-genes (those with an HI score of 0–10% or a pLI score of ≥0.9) are highlighted in bold. HI and pLI scores were derived from DECIPHER in January 2021 (https://www.deciphergenomics.org).

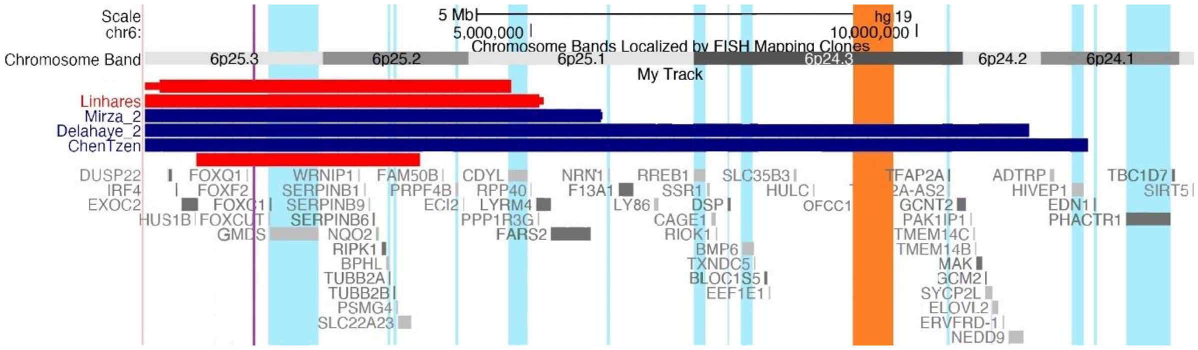


**Figure S1. Cleft lip and/or palate.** Overview of individuals with isolated cleft palate (red) and cleft lip and palate (blue). The genes *FOXF2* and *OFCC1* are indicated with a purple and an orange bar, respectively. *OFCC1* did not appear as an OMIM gene in the UCSC browser (https://genome.ucsc.edu), but it was added in the figure because of its proposed relation to orofacial clefting (see Discussion).

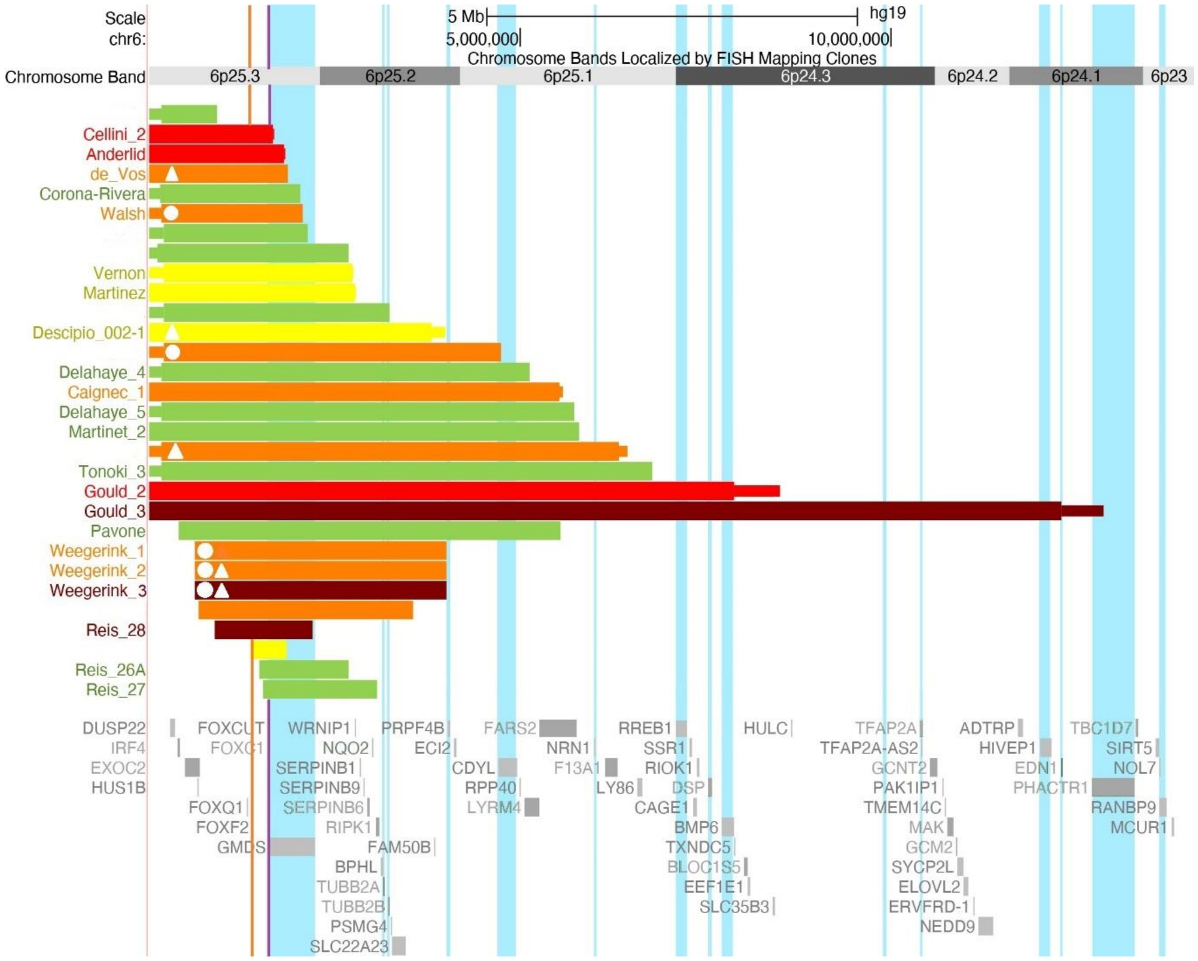


**Figure S2. Hearing impairment and middle/inner ear abnormalities.** Overview of patients for whom information on the presence and severity of hearing impairment was available: no (green), mild (yellow), moderate (orange), severe (red) and profound (dark red) hearing impairment. White circles indicate the presence of abnormalities of the tympanic membrane and/or middle ear ossicles. White triangles represent abnormalities of the cochlea and/or its nerves. For four individuals with middle/inner ear abnormalities, the severity of hearing impairment was unknown, and they are not included in this figure. The genes *FOXC1* and *FOXF2* are indicated with a purple and orange vertical lines, respectively (see Discussion).

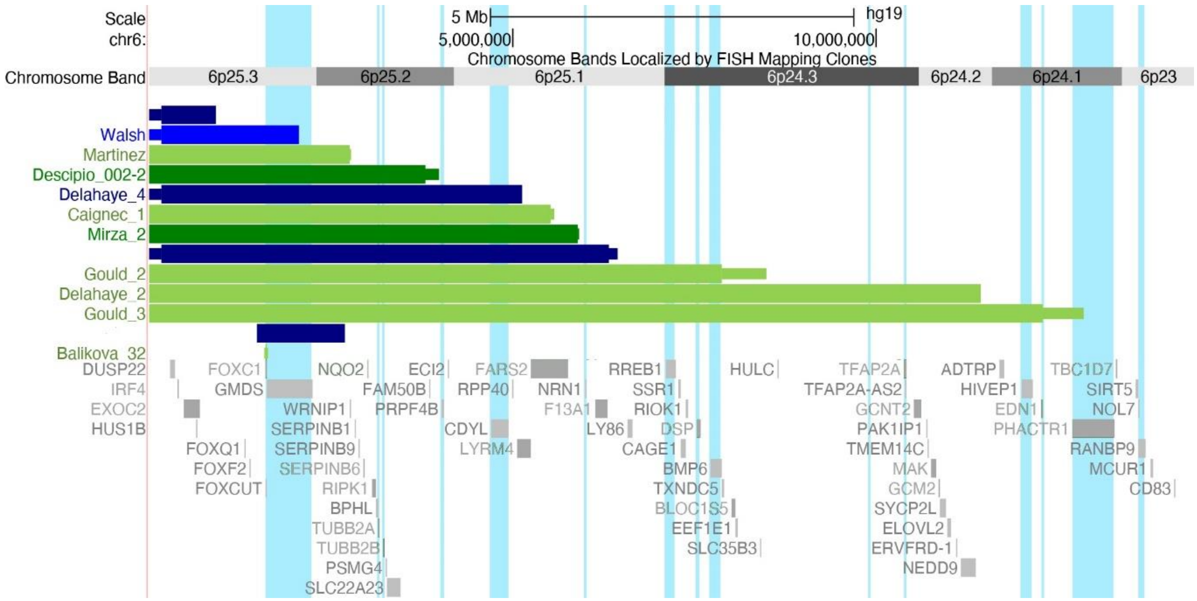


**Figure S3. Positional foot deformity and pes planus.** Overview of patients with positional foot deformities (green) and pes planus (blue). Darker colours represent the presence of hypotonia in combination with the foot deformity. Please note that in deletions with lighter colours, the presence of hypotonia was unknown.

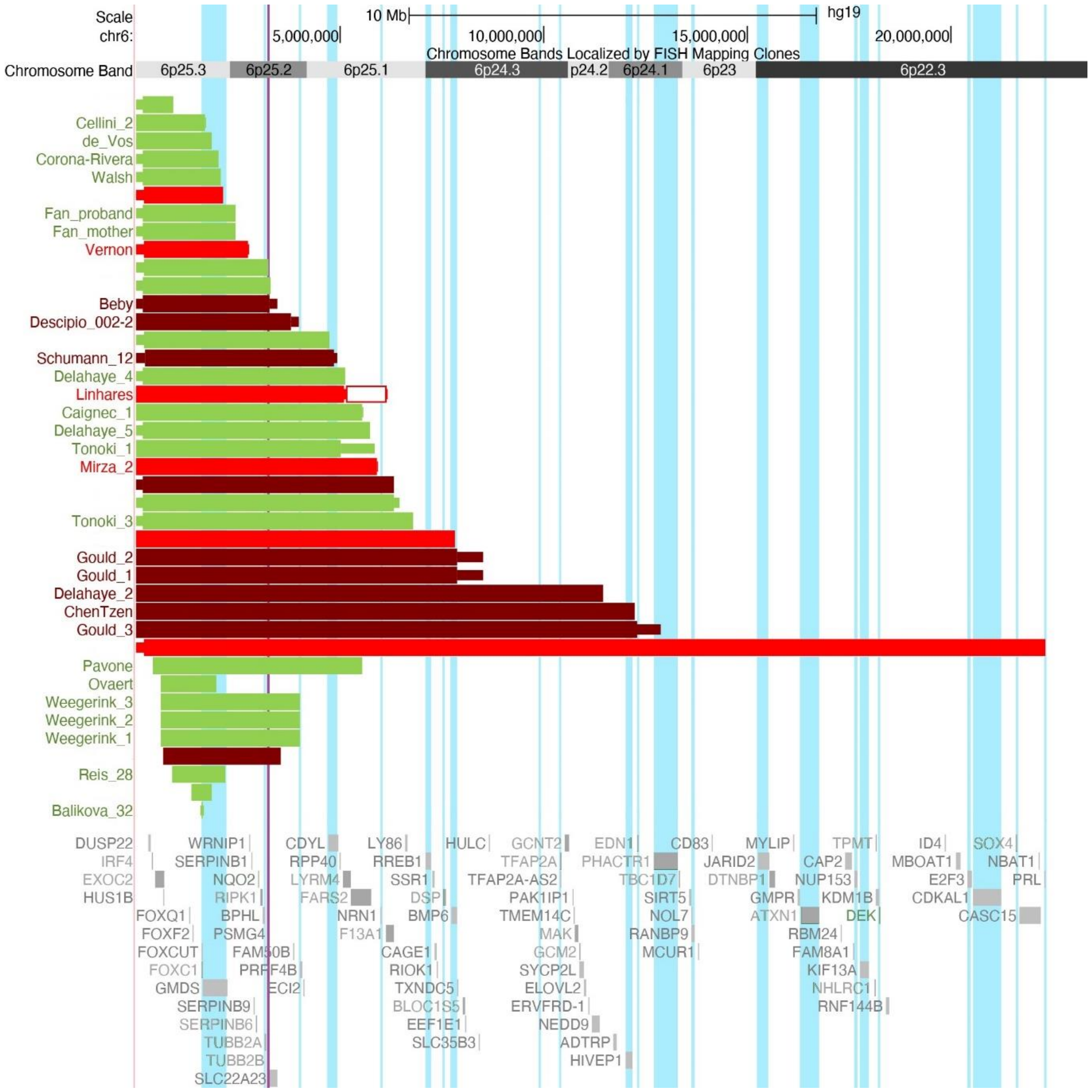


**Figure S4. Cerebellar abnormality and Dandy-Walker malformation.** Overview of individuals with hindbrain abnormalities: patients without a cerebellar abnormality (but who might have other brain abnormalities) (green), cerebellar abnormality without (or unknown) Dandy-Walker complex (red) or with Dandy-Walker malformation/variant (dark red). The gene *TUBB2B* is indicated by the purple vertical line (see Discussion).

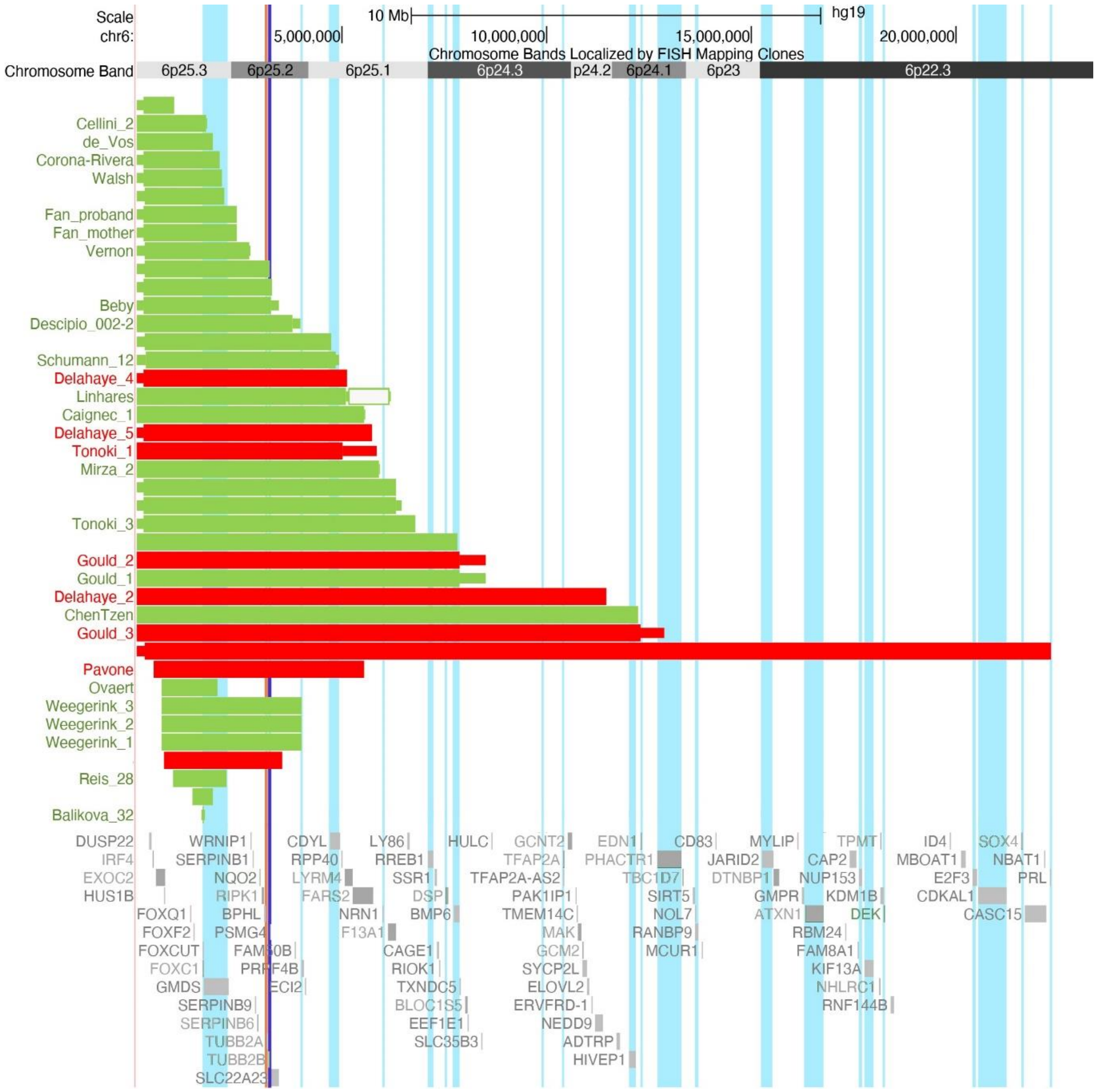


**Figure S5. Corpus callosum abnormality.** Overview of patients without corpus callosum abnormalities (but who might have other brain abnormalities) (green) and those with corpus callosum abnormalities (red). The genes *TUBB2A* and *TUBB2B* are indicated by the orange and dark blue bar, respectively (see Discussion).
