## Additional File 3 for "The phenotypic spectrum of terminal and subterminal 6p deletions based on a social media-derived cohort and literature review"

**Table S3.** Detailed overview of the phenotypic characteristics seen in individuals with (sub)terminal 6p deletions

| Phenotypic characteristic | Patient A | Subgroup B (n=14) | Subgroup C (n=5) | Subgroup D (n=13) | Subgroup E (n=7) | Total terminal (n=40) | Subterminal (n=19) | Total terminal and subterminal (n=59) | Total terminal and subterminal parents (n=13) | Total terminal and subterminal literature (n=46) |
|---|---|---|---|---|---|---|---|---|---|---|
| Parent/Literature | Parent | 2/12 | 2/3 | 3/10 | 2/5 | 10/30 | 3/16 | 13/46 | | |
| Sex (F/M) | [redacted] | 13/1 | 2/3 | 7/6 | 4/2 | 26/13 | 10/7 | 36/20 | 7/6 | 29/14 |
| **Pregnancy and neonatal period** | | | | | | | | | | |
| Intrauterine growth retardation | + | 1/6 | 0/2 | 2/8 | 2/4 | 6/21 | 2/4 | 8/25 | 4/13 | 4/12 |
| Polyhydramnios | - | 1/5 | 0/2 | 3/8 | 0/3 | 4/19 | 0/2 | 4/21 | 3/12 | 1/9 |
| Caesarean section | - | 3/7 | 1/3 | 4/7 | 1/2 | 9/20 | 3/4 | 12/24 | 6/13 | 6/11 |
| Neonatal asphyxia | + | 0/6 | 0/2 | 2/3 | 1/2 | 4/14 | 0/3 | 4/17 | 4/13 | 0/4 |
| Neonatal feeding problems | + | 2/6 | 1/2 | 3/3 | 2/2 | 9/14 | 3/3 | 12/17 | 10/13 | 2/4 |
| Prolonged neonatal jaundice | + | 0/6 | 1/2 | 1/3 | 1/2 | 4/14 | 1/3 | 5/17 | 5/13 | 0/4 |
| Temperature dysregulation necessitating incubation | + | 0/6 | 0/2 | 2/3 | 0/2 | 3/14 | 0/3 | 3/17 | 3/13 | 0/4 |
| Hospitalization after birth | + | 0/3 | 1/2 | 3/3 | 2/2 | 7/11 | 1/2 | 8/13 | 8/12 | 0/1 |
| For congenital problems | - | 0/0 | 1/1 | 3/3 | 0/2 | 4/7 | 1/1 | 5/8 | 5/8 | 0/0 |
| Due to premature birth | + | 0/0 | 0/1 | 0/3 | 1/2 | 2/7 | 1/1 | 3/8 | 3/8 | 0/0 |
| **General** | | | | | | | | | | |
| Stature: Short/normal/long | Short | 3/7/0 | 0/4/0 | 0/10/1 | 1/1/0 | 5/22/1 | 3/5/0 | 8/27/1 | 4/8/1 | 4/19/0 |
| Head circumference: Micro-/Normo-/Macrocephaly | Micro-cephaly | 1/6/2 | 0/4/0 | 0/9/2 | 1/1/0 | 3/20/4 | 0/5/0 | 3/25/4 | 1/9/1 | 2/16/3 |
| Large fontanels | - | 1/5 | 0/1 | 1/4 | 1/2 | 3/13 | 2/2 | 5/15 | 4/10 | 1/5 |
| **Eyes and vision** | | | | | | | | | | |
| Hypertelorism | - | 9/11 | 4/5 | 12/12 | 7/7 | 32/36 | 8/8 | 40/44 | 10/13 | 30/31 |
| Abnormal lacrimal duct morphology | + | 0/1 | 0/1 | 0/3 | 0/1 | 1/7 | 0/2 | 1/9 | 1/9 | 0/0 |
| Abnormalities of vision | + | 8/9 | 4/5 | 9/9 | 2/3 | 24/27 | 7/7 | 31/34 | 11/13 | 20/21 |
| Mildly reduced visual acuity | + | 4/8 | 2/4 | 5/9 | 1/2 | 13/24 | 5/7 | 18/31 | 4/11 | 14/20 |
| Severe visual impairment/blindness | - | 1/8 | 1/4 | 1/9 | 1/2 | 4/24 | 1/7 | 5/31 | 4/11 | 1/20 |
| Amblyopia | - | 1/8 | 0/4 | 2/9 | 0/2 | 3/24 | 0/7 | 3/31 | 2/11 | 1/20 |
| Refraction problems | - | 2/4 | 3/3 | 6/7 | 1/2 | 12/17 | 4/5 | 16/22 | 6/11 | 10/11 |
| Hypermetropia | n.a. | 2/2 | 0/2 | 5/6 | 1/1 | 8/11 | 2/2 | 10/13 | 4/4 | 6/9 |
| Mild hypermetropia | n.a. | 0/2 | 0/0 | 4/5 | 1/1 | 5/8 | 2/2 | 7/10 | 2/4 | 5/6 |
| High hypermetropia | n.a. | 2/2 | 0/0 | 1/5 | 0/1 | 3/8 | 0/2 | 3/10 | 2/4 | 1/6 |
| Mild myopia | n.a. | 0/2 | 2/2 | 1/6 | 0/1 | 3/11 | 0/2 | 3/13 | 0/4 | 3/9 |
| Eye movement abnormalities | - | 6/10 | 1/3 | 7/9 | 1/4 | 15/27 | 3/5 | 18/32 | 4/13 | 14/19 |
| Strabismus | n.a. | 4/6 | 1/1 | 5/7 | 0/1 | 10/15 | 1/3 | 11/18 | 0/4 | 11/14 |
| Nystagmus | n.a. | 1/6 | 0/1 | 1/7 | 1/1 | 3/15 | 2/3 | 5/18 | 3/4 | 2/14 |
| Glaucoma | - | 4/13 | 1/3 | 1/11 | 1/7 | 7/35 | 12/19 | 19/54 | 3/12 | 16/42 |
| Cataract | - | 1/13 | 0/3 | 1/11 | 0/7 | 2/35 | 2/19 | 4/54 | 0/12 | 4/42 |
| Microphthalmos | - | 1/13 | 0/3 | 0/11 | 2/7 | 3/35 | 0/19 | 3/54 | 0/12 | 3/42 |
| A/hypoplasia of the optic nerve | - | 1/13 | 1/3 | 1/11 | 1/7 | 4/35 | 0/19 | 4/54 | 2/12 | 2/42 |
| Coloboma | - | 0/13 | 1/3 | 1/11 | 1/7 | 3/35 | 2/19 | 5/54 | 3/12 | 2/42 |
| Abnormality of retinal pigmentation | - | 0/13 | 0/3 | 0/11 | 1/7 | 1/35 | 2/19 | 3/54 | 1/12 | 2/42 |
| Anterior segment dysgenesis | - | 8/12 | 2/3 | 10/10 | 4/6 | 24/32 | 14/18 | 38/50 | 5/9 | 33/41 |
| Axenfeld and/or Rieger anomaly | n.a. | 8/8 | 2/2 | 9/10 | 3/4 | 22/24 | 14/14 | 36/38 | 4/5 | 32/33 |
| Axenfeld anomaly | n.a. | 8/8 | 2/2 | 6/9 | 2/3 | 18/22 | 11/13 | 29/35 | 1/3 | 28/32 |
| Posterior Embryotoxon | n.a. | 4/6 | 2/2 | 6/6 | 1/2 | 13/16 | 6/6 | 19/22 | 1/1 | 18/21 |
| Iris strands attached to Schwalbe's line | n.a. | 3/6 | 0/2 | 3/6 | 2/2 | 8/16 | 4/6 | 12/22 | 0/1 | 12/21 |
| Goniodysgenesis | n.a. | 3/6 | 0/2 | 0/6 | 0/2 | 3/16 | 0/6 | 3/22 | 0/1 | 3/21 |
| Rieger anomaly | n.a. | 5/8 | 1/2 | 6/9 | 2/3 | 14/22 | 9/13 | 23/35 | 2/3 | 21/32 |
| Iris hypoplasia | n.a. | 2/3 | 0/1 | 2/6 | 2/2 | 6/12 | 3/5 | 9/17 | 1/2 | 8/15 |
| Corectopia | n.a. | 2/3 | 1/1 | 4/6 | 0/2 | 7/12 | 1/4 | 8/16 | 1/2 | 7/14 |
| Polycoria | n.a. | 1/3 | 0/1 | 0/6 | 0/2 | 1/12 | 1/4 | 2/16 | 0/2 | 2/14 |
| Anisocoria | n.a. | 1/3 | 0/1 | 0/6 | 0/2 | 1/12 | 0/4 | 1/16 | 0/2 | 1/14 |
| Corneal opacity | n.a. | 2/8 | 0/2 | 2/10 | 2/4 | 6/24 | 0/14 | 6/38 | 1/5 | 5/33 |
| Megalocornea | n.a. | 1/8 | 0/2 | 0/10 | 1/4 | 2/24 | 2/14 | 4/38 | 0/5 | 4/33 |
| **Ears, hearing and balance** | | | | | | | | | | |
| Periauricular skin pits | - | 0/4 | 1/4 | 0/3 | 2/3 | 3/15 | 0/3 | 3/18 | 1/12 | 2/6 |
| Dysplastic outer ear and/or low-set ears | - | 3/7 | 0/3 | 4/12 | 6/7 | 13/30 | 2/5 | 15/35 | 1/12 | 14/23 |
| Abnormalities of middle and/or inner ear | - | 2/4 | 2/3 | 3/4 | 2/2 | 9/14 | 3/5 | 12/19 | 5/12 | 7/7 |
| Abnormality of the cochlea | n.a. | 1/2 | 1/2 | 0/3 | 1/2 | 3/9 | 2/3 | 5/12 | 1/5 | 4/7 |
| Abnormality of the tympanic membrane | n.a. | 1/2 | 0/2 | 1/3 | 0/2 | 2/9 | 2/3 | 4/12 | 1/5 | 3/7 |
| Abnormal middle ear ossicles | n.a. | 0/2 | 0/1 | 1/2 | 0/0 | 1/5 | 3/3 | 4/8 | 0/1 | 4/7 |
| Abnormality of the nerves of inner ear | n.a. | 0/2 | 0/2 | 1/3 | 0/2 | 1/9 | 2/3 | 3/12 | 1/5 | 2/7 |
| Hearing impairment | - | 9/12 | 2/3 | 7/11 | 4/4 | 22/31 | 10/13 | 32/44 | 7/11 | 25/33 |
| Mild hearing impairment | n.a. | 2/6 | 1/1 | 0/3 | 0/2 | 3/12 | 1/6 | 4/18 | 1/4 | 3/14 |
| Moderate hearing impairment | n.a. | 2/6 | 0/1 | 3/3 | 0/2 | 5/12 | 3/6 | 8/18 | 3/4 | 5/14 |

| | | | | | | | | | | |
|---|---|---|---|---|---|---|---|---|---|---|
| Severe hearing impairment | n.a. | 2/6 | 0/1 | 0/3 | 1/2 | 3/12 | 0/6 | 3/18 | 0/4 | 3/14 |
| Profound hearing impairment | n.a. | 0/6 | 0/1 | 0/3 | 1/2 | 1/12 | 2/6 | 3/18 | 0/4 | 3/14 |
| Progressive hearing impairment | n.a. | 2/7 | 1/1 | 1/3 | 0/1 | 4/12 | 3/4 | 7/16 | 3/7 | 4/9 |
| Sensorineural hearing loss | n.a. | 4/8 | 1/1 | 1/5 | 0/0 | 6/14 | 1/6 | 7/20 | 0/3 | 7/17 |
| Conductive hearing loss | n.a. | 1/8 | 0/1 | 3/5 | 0/0 | 4/14 | 0/6 | 4/20 | 1/3 | 3/17 |
| Mixed hearing loss | n.a. | 3/8 | 0/1 | 1/5 | 0/0 | 4/14 | 5/6 | 9/20 | 2/3 | 7/17 |
| Balance problems | + | 3/4 | 1/3 | 3/3 | 0/2 | 8/13 | 3/6 | 11/19 | 9/13 | 2/6 |
| Vestibular dysfunction | - | 0/3 | 0/1 | 2/3 | 0/0 | 2/8 | 0/3 | 2/11 | 2/9 | 0/2 |
| Cerebellum dysfunction | - | 1/3 | 0/1 | 0/3 | 0/0 | 1/8 | 1/3 | 2/11 | 1/9 | 1/2 |
| Balance disorder of unknown cause | + | 0/3 | 0/1 | 0/3 | 0/0 | 1/8 | 0/3 | 1/11 | 1/9 | 0/2 |
| **Oral region** | | | | | | | | | | |
| Isolated cleft palate | - | 0/7 | 0/3 | 2/10 | 0/5 | 2/26 | 1/2 | 3/28 | 2/11 | 1/17 |
| Cleft lip and cleft palate | - | 0/7 | 0/3 | 1/10 | 2/5 | 3/26 | 0/2 | 3/28 | 0/11 | 3/17 |
| Dental problems | - | 4/9 | 2/3 | 5/8 | 0/0 | 11/21 | 4/9 | 15/30 | 5/9 | 10/21 |
| Abnormal dental morphology | n.a. | 2/4 | 1/2 | 2/5 | 0/0 | 5/11 | 1/4 | 6/15 | 3/5 | 3/10 |
| Reduced number of teeth | n.a. | 2/4 | 0/2 | 1/5 | 0/0 | 3/11 | 2/4 | 5/15 | 1/5 | 4/10 |
| **Internal organs** | | | | | | | | | | |
| Gastroesophageal reflux | - | 0/2 | 0/2 | 1/3 | 1/2 | 2/10 | 2/3 | 4/13 | 4/13 | 0/0 |
| Requiring medication or surgery | n.a. | 0/0 | 0/0 | 1/1 | 1/1 | 2/2 | 1/2 | 3/4 | 3/4 | 0/0 |
| Feeding difficulties | - | 1/4 | 2/3 | 3/3 | 2/2 | 8/13 | 2/3 | 10/16 | 8/13 | 2/3 |
| Requiring tube feeding | n.a. | 0/1 | 0/2 | 1/3 | 2/2 | 3/8 | 2/2 | 5/10 | 5/8 | 0/2 |
| Lower intestinal problems | - | 1/3 | 0/2 | 2/3 | 1/2 | 4/11 | 1/3 | 5/14 | 4/13 | 1/1 |
| Bowel incontinence | n.a. | 1/1 | 0/0 | 1/2 | 0/1 | 2/4 | 1/1 | 3/5 | 2/4 | 1/1 |
| Constipation | n.a. | 0/1 | 0/0 | 2/2 | 1/1 | 3/4 | 1/1 | 4/5 | 4/4 | 0/1 |
| Congenital heart defect | + | 3/11 | 2/3 | 6/11 | 4/5 | 16/31 | 6/11 | 22/42 | 7/13 | 15/29 |
| Atrial septal defect | - | 2/3 | 0/2 | 3/6 | 1/4 | 6/16 | 4/6 | 10/22 | 1/7 | 9/15 |
| Ventricular septal defect | + | 0/3 | 0/2 | 0/6 | 3/4 | 4/16 | 0/6 | 4/22 | 2/7 | 2/15 |
| Patent foramen ovale | - | 0/3 | 1/2 | 1/6 | 2/4 | 4/16 | 1/6 | 5/22 | 0/7 | 5/15 |
| Patent ductus arteriosus | - | 1/3 | 0/2 | 1/6 | 1/4 | 3/16 | 0/6 | 3/22 | 0/7 | 3/15 |
| Abnormality of heart valves | - | 1/3 | 2/2 | 1/6 | 1/4 | 5/16 | 4/6 | 9/22 | 2/7 | 7/15 |
| Abnormal tricuspid valve | n.a. | 0/1 | 1/2 | 0/1 | 0/1 | 2/5 | 1/4 | 3/9 | 0/2 | 3/7 |
| Pulmonic stenosis | n.a. | 0/1 | 0/2 | 1/1 | 0/1 | 1/5 | 2/4 | 3/9 | 0/2 | 3/7 |
| Bicuspid aortic valve | n.a. | 0/1 | 1/2 | 0/1 | 0/1 | 1/5 | 1/4 | 2/9 | 2/2 | 0/7 |
| Aortic valve stenosis | n.a. | 0/1 | 0/2 | 0/1 | 0/1 | 0/5 | 2/4 | 2/9 | 0/2 | 2/7 |
| Coarctation of aorta | - | 0/3 | 0/2 | 0/6 | 1/4 | 1/16 | 0/6 | 1/22 | 1/7 | 0/15 |
| Hypoplastic left heart | - | 0/3 | 0/2 | 0/6 | 1/4 | 1/16 | 0/6 | 1/22 | 0/7 | 1/15 |
| Kidney abnormalities | - | 1/3 | 0/2 | 1/3 | 3/4 | 5/13 | 1/5 | 6/18 | 2/13 | 4/5 |
| Hydronephrosis | n.a. | 0/1 | 0/0 | 0/1 | 1/3 | 1/5 | 0/1 | 1/6 | 0/2 | 1/4 |
| Renal cysts | n.a. | 0/1 | 0/0 | 0/1 | 2/3 | 2/5 | 0/1 | 2/6 | 1/2 | 1/4 |
| Renal hypoplasia | n.a. | 0/1 | 0/0 | 1/1 | 0/3 | 1/5 | 0/1 | 1/6 | 1/2 | 0/4 |
| Vesicoureteral reflux | n.a. | 1/1 | 0/0 | 0/1 | 0/3 | 1/5 | 0/1 | 1/6 | 0/2 | 1/4 |
| Abnormalities of male genitalia | + | 1/1 | 0/2 | 1/5 | 1/2 | 4/11 | 1/3 | 5/14 | 2/7 | 3/7 |
| Micropenis | - | 1/1 | 0/0 | 1/1 | 1/1 | 3/4 | 1/1 | 4/5 | 1/2 | 3/3 |
| **Immune and endocrine system** | | | | | | | | | | |
| Recurrent infections | + | 3/3 | 2/3 | 2/4 | 0/2 | 8/13 | 4/4 | 12/17 | 7/12 | 5/5 |
| Recurrent ear infections | + | 3/3 | 2/2 | 2/2 | 0/0 | 8/8 | 2/4 | 10/12 | 5/7 | 5/5 |
| Recurrent lower respiratory tract infections | + | 1/3 | 1/2 | 1/2 | 0/0 | 4/8 | 1/4 | 5/12 | 5/7 | 0/5 |
| Recurrent upper respiratory tract infections | + | 3/3 | 0/2 | 1/2 | 0/0 | 5/8 | 0/4 | 5/12 | 3/7 | 2/5 |
| Ear tubes placed | - | 1/2 | 2/3 | 1/3 | 1/2 | 5/11 | 3/5 | 8/16 | 5/12 | 3/4 |
| Hypothyroidism | + | 0/5 | 0/2 | 1/6 | 0/2 | 2/16 | 0/5 | 2/21 | 1/13 | 1/8 |
| **Trunk and extremities** | | | | | | | | | | |
| Umbilical hernia | - | 1/3 | 2/4 | 0/7 | 1/1 | 4/17 | 0/4 | 4/21 | 1/13 | 3/8 |
| Abnormalities of vertebral column | - | 2/4 | 0/3 | 2/4 | 2/4 | 6/16 | 1/3 | 7/19 | 1/11 | 6/8 |
| Scoliosis | n.a. | 2/2 | 0/0 | 1/2 | 2/2 | 5/6 | 1/1 | 6/7 | 0/1 | 6/6 |
| Kyphosis | n.a. | 0/2 | 0/0 | 1/2 | 0/2 | 1/6 | 1/1 | 2/7 | 1/1 | 1/6 |
| Abnormal vertebral morphology | - | 1/4 | 0/2 | 1/4 | 0/2 | 2/13 | 1/3 | 3/16 | 0/12 | 3/4 |
| Hip dysplasia | u | 0/3 | 0/2 | 2/5 | 0/1 | 2/11 | 3/4 | 5/15 | 3/10 | 2/5 |
| Spina bifida | - | 0/2 | 0/4 | 1/3 | 0/2 | 1/12 | 0/4 | 1/16 | 1/13 | 0/3 |
| Joint hypermobility | - | 2/3 | 0/2 | 3/4 | 1/3 | 6/13 | 4/4 | 10/17 | 7/13 | 3/4 |
| Finger joint hypermobility | n.a. | 0/2 | 0/0 | 2/3 | 0/1 | 2/6 | 0/4 | 2/10 | 2/7 | 0/3 |
| Generalized joint laxity | n.a. | 1/2 | 0/0 | 0/3 | 0/1 | 1/6 | 2/4 | 3/10 | 3/7 | 0/3 |
| Hyperextensibility at elbow | n.a. | 0/2 | 0/0 | 1/3 | 0/1 | 1/6 | 0/4 | 1/10 | 0/7 | 1/3 |
| Knee joint hypermobility | n.a. | 0/2 | 0/0 | 1/3 | 0/1 | 1/6 | 0/4 | 1/10 | 0/7 | 1/3 |
| Flexion contracture | - | 0/2 | 0/4 | 2/3 | 0/3 | 2/13 | 0/2 | 2/15 | 2/11 | 0/4 |
| Pes planus/positional foot deformity | + | 2/10 | 1/4 | 4/8 | 3/6 | 11/29 | 2/6 | 13/35 | 3/13 | 10/22 |
| Bone deformities (epiphyseal) | u | 5/5 | 0/1 | 5/5 | 1/1 | 11/12 | 2/4 | 13/16 | 3/5 | 10/11 |
| Femoral bone deformity | n.a. | 2/5 | 0/0 | 1/5 | 1/1 | 4/11 | 0/2 | 4/13 | 0/3 | 4/10 |
| Ulnar bone deformity | n.a. | 2/5 | 0/0 | 0/5 | 0/1 | 2/11 | 0/2 | 2/13 | 0/3 | 2/10 |
| Humerus bone deformity | n.a. | 1/5 | 0/0 | 1/5 | 1/1 | 3/11 | 0/2 | 3/13 | 1/3 | 2/10 |
| Delayed ossification | n.a. | 0/5 | 0/0 | 2/5 | 1/1 | 3/11 | 0/2 | 3/13 | 1/3 | 2/10 |
| **Nervous system** | | | | | | | | | | |
| Brain abnormalities on imaging | + | 7/8 | 4/4 | 9/11 | 7/7 | 28/31 | 8/9 | 36/40 | 10/11 | 26/29 |
| Abnormal cortical gyration | - | 0/7 | 0/4 | 0/9 | 0/7 | 0/28 | 1/8 | 1/36 | 1/10 | 0/26 |

| | | | | | | | | | | |
|---|---|---|---|---|---|---|---|---|---|---|
| Abnormality of cerebellum | - | 2/7 | 2/4 | 4/9 | 7/7 | 15/28 | 1/8 | 16/36 | 5/10 | 11/26 |
| Dandy-Walker complex | n.a. | 0/1 | 2/2 | 2/4 | 5/5 | 9/12 | 1/1 | 10/13 | 2/2 | 8/11 |
| Abnormality of cerebral vasculature | - | 1/7 | 0/4 | 1/9 | 0/7 | 2/28 | 1/8 | 3/36 | 1/10 | 2/26 |
| Aplasia/hypoplasia of cerebrum | - | 1/7 | 0/4 | 1/9 | 0/7 | 2/28 | 0/8 | 2/36 | 1/10 | 1/26 |
| Corpus callosum abnormality | - | 0/7 | 0/4 | 3/9 | 4/7 | 7/28 | 2/8 | 9/36 | 2/10 | 7/26 |
| Cortical dysplasia/ migration disorder | - | 1/7 | 0/4 | 0/9 | 0/7 | 1/28 | 0/8 | 1/36 | 0/10 | 1/26 |
| Delayed myelination | - | 1/7 | 0/4 | 1/9 | 0/7 | 2/28 | 0/8 | 2/36 | 1/10 | 1/26 |
| Intracranial cystic lesion | - | 0/7 | 0/4 | 0/9 | 0/7 | 0/28 | 2/8 | 2/36 | 1/10 | 1/26 |
| Intracranial haemorrhage | - | 0/7 | 1/4 | 0/9 | 0/7 | 1/28 | 0/8 | 1/36 | 1/10 | 0/26 |
| Ventriculomegaly/hydrocephalus | - | 3/8 | 3/4 | 5/9 | 5/7 | 16/29 | 3/8 | 19/37 | 3/10 | 16/27 |
| Dilated perivascular (VR) spaces | u | 2/6 | 0/4 | 1/9 | 0/5 | 3/24 | 2/7 | 5/31 | 2/6 | 3/25 |
| White matter abnormalities | u | 5/6 | 0/4 | 5/9 | 0/5 | 10/24 | 6/7 | 16/31 | 4/6 | 12/25 |
| Abnormality of the basal ganglia | u | 2/6 | 0/4 | 1/9 | 0/5 | 3/24 | 0/7 | 3/31 | 1/6 | 2/25 |
| Seizures | - | 1/4 | 1/2 | 2/4 | 1/3 | 5/14 | 1/2 | 6/16 | 3/11 | 3/5 |
| Absence seizures | n.a. | 0/1 | 1/1 | 0/2 | 0/1 | 1/5 | 1/1 | 2/6 | 2/3 | 0/3 |
| Febrile seizures | n.a. | 1/1 | 0/1 | 1/2 | 0/1 | 2/5 | 1/1 | 3/6 | 2/3 | 1/3 |
| Generalized-onset seizures | n.a. | 0/1 | 0/1 | 1/2 | 0/1 | 1/5 | 1/1 | 2/6 | 1/3 | 1/3 |
| Hypotonia | + | 3/3 | 4/4 | 7/9 | 2/2 | 17/19 | 3/5 | 20/24 | 10/10 | 10/14 |
| Torticollis | - | 0/1 | 1/3 | 1/2 | 0/2 | 2/9 | 1/1 | 3/10 | 3/9 | 0/1 |
| Abnormal pain sensation | - | 0/1 | 0/2 | 3/3 | 0/2 | 3/9 | 1/2 | 4/11 | 4/11 | 0/0 |
| Impaired pain sensation | n.a. | 0/0 | 0/0 | 1/3 | 0/0 | 1/3 | 1/1 | 2/4 | 2/4 | 0/0 |
| Over-reactivity of the palms and/or soles | n.a. | 0/0 | 0/0 | 2/3 | 0/0 | 2/3 | 0/1 | 2/4 | 2/4 | 0/0 |
| Continuous abnormality of pain sensation | n.a. | 0/0 | 0/0 | 3/3 | 0/0 | 3/3 | 1/1 | 4/4 | 4/4 | 0/0 |
| Sensory impairment | + | 0/1 | 1/2 | 2/4 | 1/2 | 5/10 | 0/2 | 5/12 | 4/11 | 1/1 |
| **Development and behaviour** | | | | | | | | | | |
| Developmental delay | - | 11/14 | 2/3 | 10/10 | 1/1 | 24/29 | 10/12 | 34/41 | 7/10 | 27/31 |
| Sleeping problems | + | 0/2 | 0/2 | 2/3 | 0/2 | 3/10 | 2/2 | 5/12 | 5/12 | 0/0 |
| Insomnia | + | 0/0 | 0/2 | 2/2 | 0/0 | 3/3 | 2/2 | 5/5 | 5/5 | 0/0 |
| Parasomnia | + | 0/0 | 0/2 | 1/2 | 0/0 | 2/3 | 1/2 | 3/5 | 3/5 | 0/0 |
| Sleep apnoea | - | 0/0 | 0/3 | 1/4 | 1/1 | 2/9 | 2/3 | 4/12 | 4/9 | 0/3 |
| Social behaviour | + | 2/4 | 2/3 | 2/6 | 0/2 | 7/16 | 2/4 | 9/20 | 8/13 | 1/7 |
| Quiet behaviour | - | 0/4 | 1/3 | 3/6 | 1/2 | 5/16 | 1/4 | 6/20 | 6/13 | 0/7 |
| Aggressive behaviour | - | 0/4 | 0/3 | 2/6 | 0/2 | 2/16 | 1/4 | 3/20 | 0/13 | 3/7 |
| Hyperactivity | - | 1/4 | 1/3 | 0/6 | 0/2 | 2/16 | 1/4 | 3/20 | 2/13 | 1/7 |
| Shyness | - | 0/4 | 1/3 | 1/6 | 0/2 | 2/16 | 1/4 | 3/20 | 3/13 | 0/7 |
| Impaired social interactions | - | 0/4 | 1/3 | 0/6 | 0/2 | 1/16 | 1/4 | 2/20 | 1/13 | 1/7 |
| Attention-deficit hyperactivity disorder (ADHD) | - | 0/2 | 1/3 | 0/4 | 0/2 | 1/12 | 0/4 | 1/16 | 1/12 | 0/4 |
| Autistic behaviour/disorder | - | 1/2 | 2/3 | 1/4 | 0/2 | 4/12 | 2/4 | 6/16 | 2/12 | 4/4 |
